# Beneficial reconstitution of gut microbiota and control of alpha-synuclein and curli-amyloids-producing enterobacteria, by beta 1,3-1,6 glucans in a clinical pilot study of autism and potentials in neurodegenerative diseases

**DOI:** 10.1101/2021.10.26.21265505

**Authors:** Kadalraja Raghavan, Vidyasagar Devaprasad Dedeepiya, Naoki Yamamoto, Nobunao Ikewaki, Tohru Sonoda, Masaru Iwasaki, Ramesh Shankar Kandaswamy, Rajappa Senthilkumar, Senthilkumar Preethy, Samuel JK Abraham

## Abstract

**Background/objective:** Gut dysbiosis is one of the major pathologies in children with autism spectrum disorder (ASD). In previous studies, Aureobasidium pullulans (i.e., black yeast AFO-202-produced beta glucan found in Nichi Glucan) yielded beneficial clinical outcomes related to sleep and behaviour. Evaluation of gut microbiota of the subjects in the present randomized pilot clinical study was undertaken and compared with an aim of gaining a mechanistic insight.

**Methods:** The study involved 18 subjects with ASD who were randomly allocated: six subjects in the control group (Group 1) underwent conventional treatment comprising remedial behavioural therapies and L-carnosine 500 mg per day, and 12 subjects (Group 2) underwent supplementation with Nichi Glucan 0.5 g twice daily along with the conventional treatment for 90 days. The subjects’ stool samples were collected at baseline and after the intervention. Whole genome metagenome (WGM) sequencing was performed.

**Results:** WGM sequencing followed by bioinformatic analysis in 13 subjects who completed the study showed that among genera of relevance, the abundance of Enterobacteria was decreased almost to zero in Group 2 after intervention, whereas it increased from 0.36% to 0.85% in Group 1. The abundance of Bacteroides increased from 16.84% to 19.09% in Group 1, whereas it decreased from 11.60% to 11.43% in Group 2. The abundance of Prevotella increased in both Group 1 and Group 2. The decrease in abundance of lactobacillus was significant in Group 2 compared to Group 1. Among species, a decrease was seen in *Escherichia coli, Akkermansia muciniphila CAG:154, Blautia spp*., *Coprobacillus sp*., and *Clostridium bolteae CAG:59*, with an increase of *Faecalibacterium prausnitzii* and *Prevotella copri*, which are both beneficial.

**Conclusion:** AFO-202 beta 1,3-1,6 glucan was able to balance the gut microbiome, which is considered beneficial in children with ASD. Effective control of curli-producing enterobacteria that leads to α-synuclein (αSyn) misfolding and accumulation, which apart from being advantageous in alleviating ASD symptoms, may have a prophylactic role in Parkinson’s and Alzheimer’s diseases where the αSyn misfolding and amyloid deposition are central to their pathogenesis. Additionally, stimulation of natural killer cells to help clear accumulated αSyn amyloids, beneficial microbiome reconstitution, and microglial rejuvenation lead us to recommend larger clinical studies in neurodevelopmental and neurodegenerative diseases of this safety-proven food supplement.

**Graphical Abstract:** The above illustration explains, stepwise, the pathogenesis as well as the way beta glucan tackles each stage of the disease process: (A) & (B) Enterobacteria secretion of curli that causes misfolding of α-synuclein (αSyn); its aggregation in enteric neuronal cells is tackled by (1) control of enterobacteria, (2) scavenging of the accumulated amyloids by activated natural killer cells, and (3) reconstitution of beneficial microbiome. (C) The prion like propagation may not occur because the accumulation of curli proteins and amyloids is controlled at the level of production and aggregation (1) as well as clearing of already accumulated deposits (3). (D) Deposition of Lewy bodies, amyloid fibrils, and misfolded αSyn are tackled by (4) microglial-based scavenging.

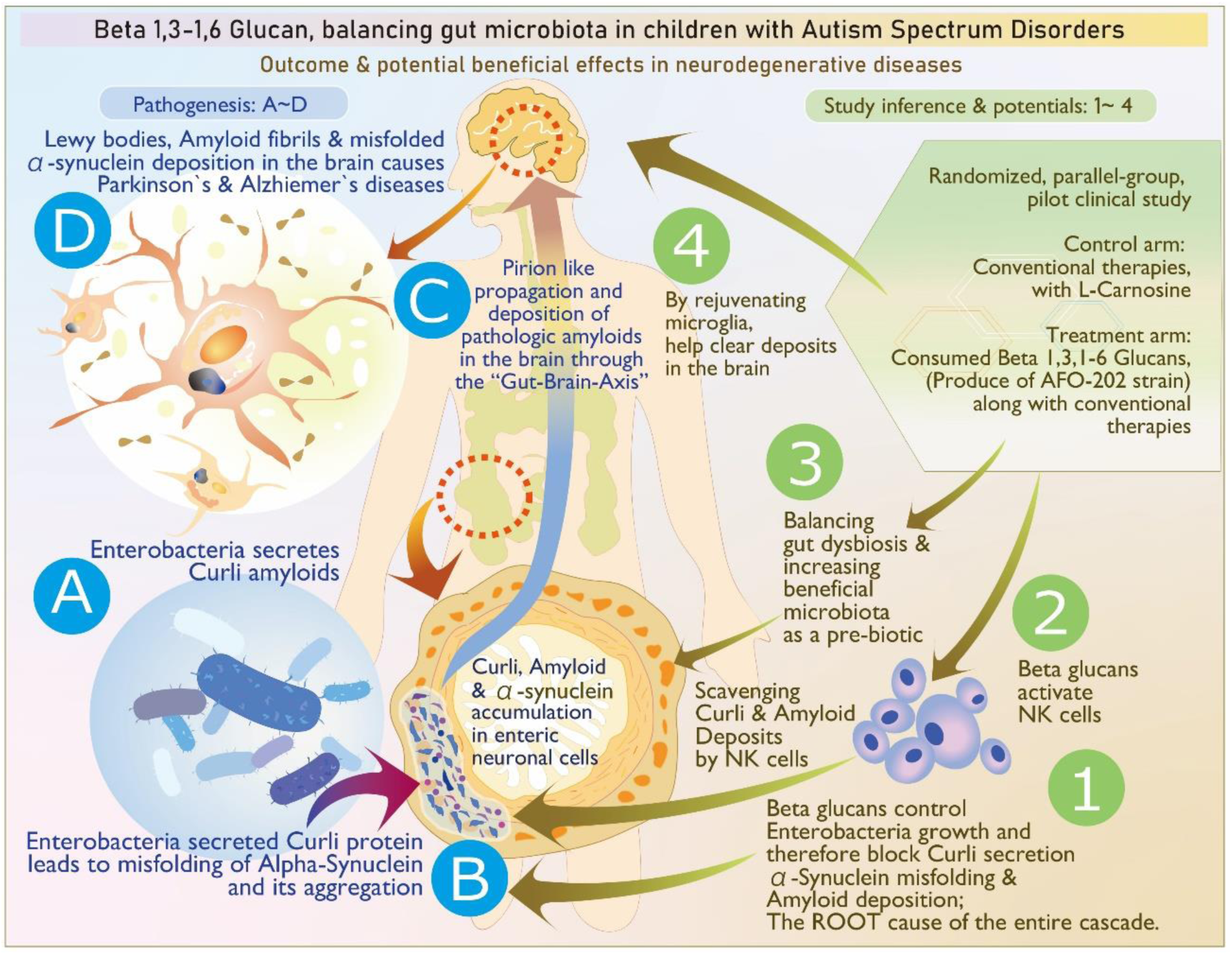

## Introduction

The emerging evidence of constant communication and interaction between the gut and the brain through the gut–brain axis has begun to unravel its significance associated with the health of the central nervous system. Any dysbiosis of the gut microbiota has been shown to influence the development and progression of neurological pathologies of developmental (autism spectrum disorder [ASD]), inflammatory (multiple sclerosis [MS]), and degenerative (Alzheimer’s disease [AD] and Parkinson’s disease [PD]) disorders [1]. The mechanisms involve activation of the immune system; production of inflammatory cytokines and chemokines (e.g., IL-6 and TNF-α); and alteration of the gut barrier permeability, which in turn is due to the increased levels of circulating lipopolysaccharide in these neurological disorders. These mechanisms modulate the neurotrophic factors, activity of the central and peripheral nervous system, and the endocrine pathways, all of which contribute to the onset or the phenotypic expression of neuropsychiatric and neurodevelopmental disorders [2]. Another important player is the amyloid protein, which has self-aggregation properties. Even non-identical amyloid proteins can accelerate reciprocal amyloid aggregation in a prion-like fashion. Nearly 30 amyloidogenic proteins are encoded by humans, whereas some functional amyloids are produced by the gut microbiome [3]. Of importance are cell-surface amyloid proteins called *curli*, which are produced by certain enterobacteria that in turn accelerate formation of α-synuclein (αSyn), which is a presynaptic neurotransmitter that is crucial in the initiation and pathogenesis of neurological disorders such as ASD, PD, AD, and MS [4]. Though these reports [2-4] discuss the correlation of the levels of these amyloids, enteric bacteria, and neural diseases, there has been no simple and safe intervention with subjective and objective correlation to a clinical benefit that can be derived based on an associated balancing of the gut microbiota. We herein report the outcome of beneficial reconstitution of gut microbiota, especially those of the bacteria associated with αSyn and curli amyloids after consumption of beta 1,3-1,6 glucans in children with ASD in a clinical pilot study. The beta-glucan studied (Nichi Glucan) was obtained from the AFO-202 strain of a black yeast called Aureobasidium pullulans that has beneficial advantages in metabolic disorders by alleviating glucotoxicity [5], lipotoxicity [6,7], lipidemia-induced hepatic fibrosis [8], inflammation [8] apart from immune-enhancement [9], and modulation in COVID-19 [10], which have been reported in translational and clinical studies. The effects of this AFO-202 beta glucan in terms of behavioural [11] and sleep pattern improvement [12], increase in levels of plasma αSyn [11], and serum melatonin [12] levels in children with ASD has been reported. We report the effects of AFO-202 beta glucan on the gut microbiome in children with ASD who participated in our pilot study.

## Methods

This study was approved by the institutional ethics committee of the hospital in which the study took place and was registered as a clinical trial in the national clinical trial registry. The caregivers of each subject gave their informed consent for inclusion before participation in the study. The study was conducted in accordance with the Declaration of Helsinki.

### Study Design

The subjects enrolled in the study had received a diagnosis of ASD from a developmental paediatrician, which was verified by a psychologist using a clinical interview for a behavioural pattern that incorporated the Childhood Autism Rating Scale score. Eighteen subjects with ASD were enrolled in this prospective, open-label, pilot clinical trial comprised of two arms. The CONSORT flow diagram is presented in Figure 1.

**Figure 1:**
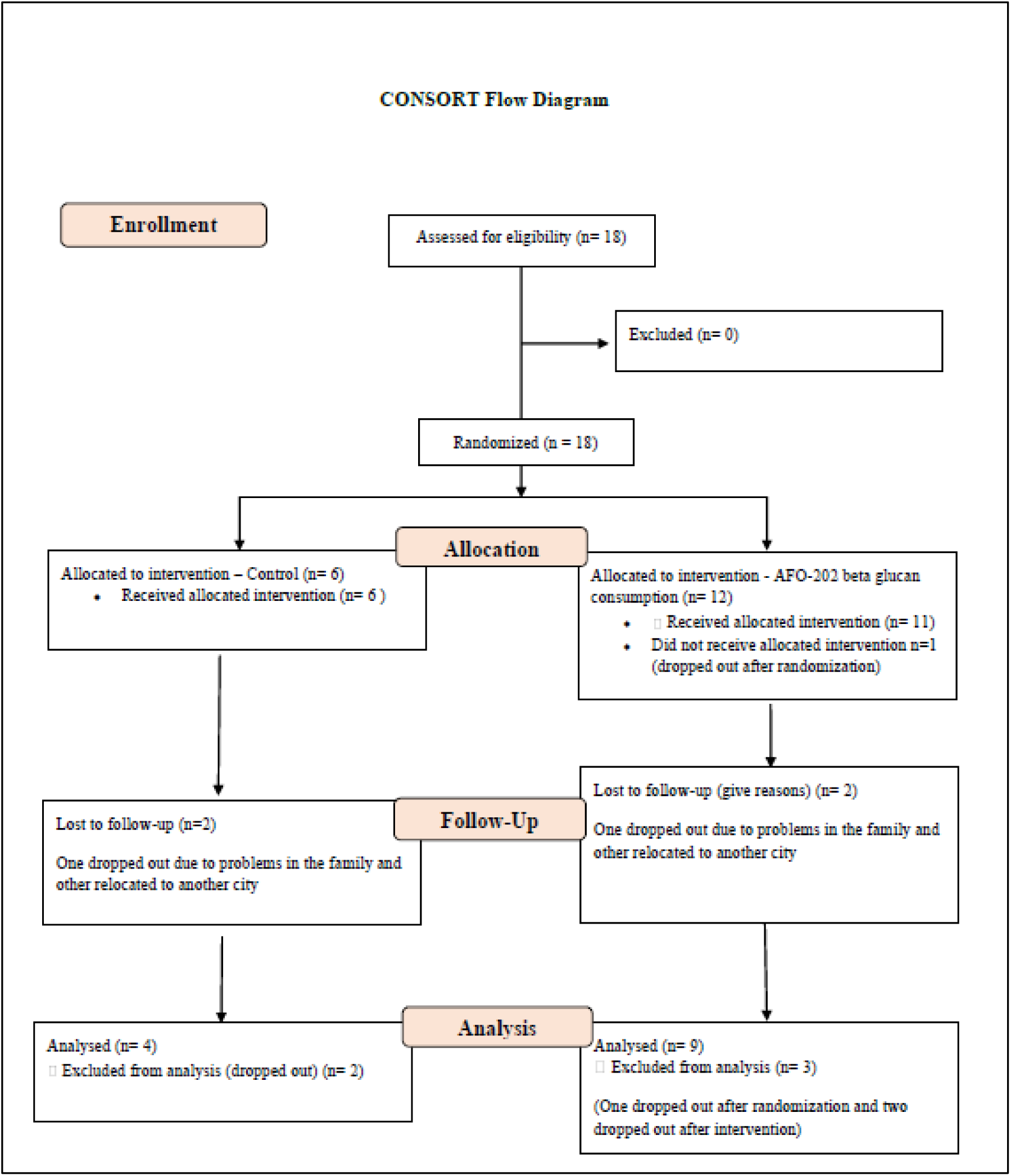
CONSORT flow diagram of the trial.

### Study Groups

Arm 1 or Group 1 (control group): Six subjects with ASD underwent conventional treatment comprising remedial behavioural therapies and L-carnosine 500 mg per day. Arm 2 or Group 2 (Nichi Glucan group): 12 subjects underwent supplementation with Nichi Glucan food supplement along with conventional treatment (remedial behavioural therapies and L-carnosine 500 mg per day). Each subject consumed two sachets (0.5 g each) of Nichi Glucan daily—one sachet with a meal twice daily—for 90 days.

The inclusion and exclusion criteria along with the assessment of behavioural and sleep pattern apart from evaluation of levels of αSyn and melatonin are available in the results of the clinical trial reported earlier [11,12].

### Faecal Sample Collection and Preparation

Faecal samples were collected at baseline and 90 days after the intervention using a sterile faecal collection kit and the samples were kept at −20 °C until they were transferred to the laboratory and processed. Samples for DNA extraction were stored at −80 °C until needed for analysis.

### DNA Extraction

Total microbial DNA was extracted from faeces of each specimen using the QIAAmp DNA Mini Kit (Qiagen) according to manufacturer’s instructions. Each batch of specimens were extracted with negative buffer control (extraction control).

### Library Preparation

Whole-genome metagenome sequencing libraries were prepared. In brief, the DNA was sheared using a Covaris ultrasonicator. Sheared DNA was subjected to a sequence of enzymatic steps for repairing the ends and tailing with dA by ligation of indexed adapter sequences. These adapter-ligated fragments were then cleaned up using SPRI beads. Next, the clean fragments are indexed using limited cycle PCR to enrich the adapter-ligated molecules. Finally, the amplified products were purified and checked before sequencing.

### Metagenome Sequencing

Prepared libraries were sequenced using Novaseq 6000 with a read length of 151 bp. The samples were taken for whole genome metagenome analysis. Initially, the reads were filtered for human DNA contamination. The alignment to the human genome was around 18.98%. The filtered reads were then aligned to bacterial, fungal, viral, and archea genomes. The overall alignment to the bacterial genome was around 40%. However, the alignment to the viral, fungi, and archaea genomes was around 0.05–0.2%. De novo assembly was carried out using the preprocessed reads to obtain the scaffolds, which were then used for gene prediction. The abundances at the phylum, genus, and species level were evaluated.

### Microbiome Bioinformatics

The following bioinformatics pipeline was used to perform whole-genome-sequencing metagenomic analysis. The quality of the raw data was analysed and the adapters were trimmed. The low-processed reads were first aligned to human genome to remove unaligned reads that were then assembled using METASPADES de novo assembler for metagenomics. After assembly, the gene prediction was performed using PRODIGAL. The predicted genes were then searched against existing genes in the NCBI database using the DIAMOND MEGAN5 program. The occurrence of dominant microbial population was studied at various levels (phylum, class, order, family, and genus) based on the taxonomic abundance in the given samples. The dominance was calculated based on the amount of sequence obtained from samples, community composition, and the contig size distribution. Chimeric sequences were identified and filtered from the analysis.

### Statistical Analysis

Statistical data were analysed using Microsoft Excel statistics package analysis software. Paired *t* tests were also calculated using this package, and *P* values < 0.05 were considered significant.

## Results

Eighteen patients who fulfilled all the selection criteria and none of the exclusion criteria were selected to begin the study. During enrolment, one participant in the treatment group (Group 2) dropped out before the study began. During the study, four subjects were lost to follow-up: two in Group 1 (one dropped out due to social problems in the family, and the other relocated to another city) and two in Group 2 (one dropped out due to social problems in the family, and the other relocated to another city). After excluding these four subjects, 13 subjects were included in the analysis.

The pre-processed reads were first aligned with the human genome (hg19) using BWA-MEM aligner to remove human genome contamination from the samples. The uncontaminated sequences were then taken for further alignment with known bacteria, fungi, virus, and archaea bacteria genomes using BWA MEM aligner. Around 7–12% of the reads mapped to the human genome, and the bacterial genome with 30–60% mapped reads.

Bacterial kingdom was the most abundant organism type one. In both Group 1 (control) and Group 2 (Nichi Glucan), both before and after intervention, phylum Firmicutes was the most abundant followed by Bacteroidetes. Although in Group 1 Proteobacteria was the next most abundant followed by Actinobacteria, this relationship was reversed in Group 2 (Figure 2).

**Figure 2:**
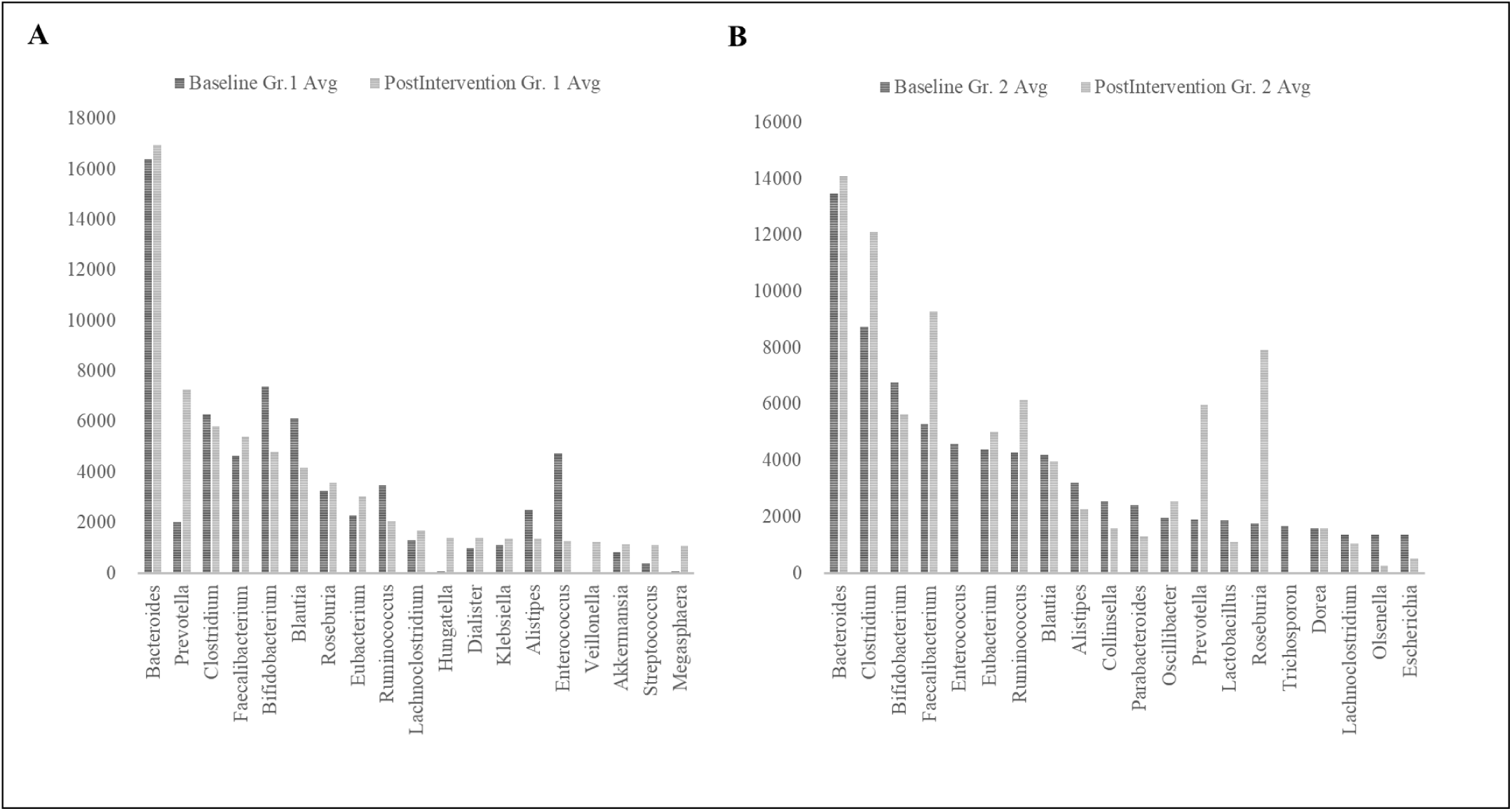
Genus Abundance of major genera identified (A) Group (Gr.) 1 at baseline versus postintervention and (B) Group (Gr.) 2 at baseline versus postintervention.

Among the genera of relevance, the abundance of Enterobacter was decreased to almost zero in Group 2 after intervention, while it increased from 0.36% to 0.85% in Group 1 (Figure 3A). The abundance of Bacteroides increased from 16.84% to 19.09% (p-value= 0.42) in Group 1, while it showed little significant difference (11.60% and 11.43%) (p value = 0.46) (Figure 3B) after intervention. The abundance of Prevotella increased in both Group 1 and Group 2 (Figure 3C). The decrease in abundance of lactobacillus was significant in Group 2 compared to Group 1 (Figure 3D). Desulfovibrio decreased from 0.40% to 0.28% in Gr.2. Species abundance increased in Group 2 after intervention. Faecalibacterium prausnitzii, Bifidobacterium longum, and Firmicutes bacterium CAG:124 represented the most abundant species (Figure 4). Escherichia coli decreased in both Group 1 and Group 2 but the difference was significant in Group 2 (p-value = 0.02) (Figure 5A). Faecalibacterium prausnitzii increased in both Group 1 and Group 2 but the difference was significant in Group 2 (p-value =0.041) (Figure 5B). Akkermansia muciniphila CAG:154 (Figure 5C) and Clostridium bolteae CAG:59 increased in the Group 1, whereas it decreased in Group 2 (Figure 5D). Prevotella copri increased in both the groups. Blautia spp., Coprobacillus sp., and several Clostridium spp. decreased in both the groups.

**Figure 3:**
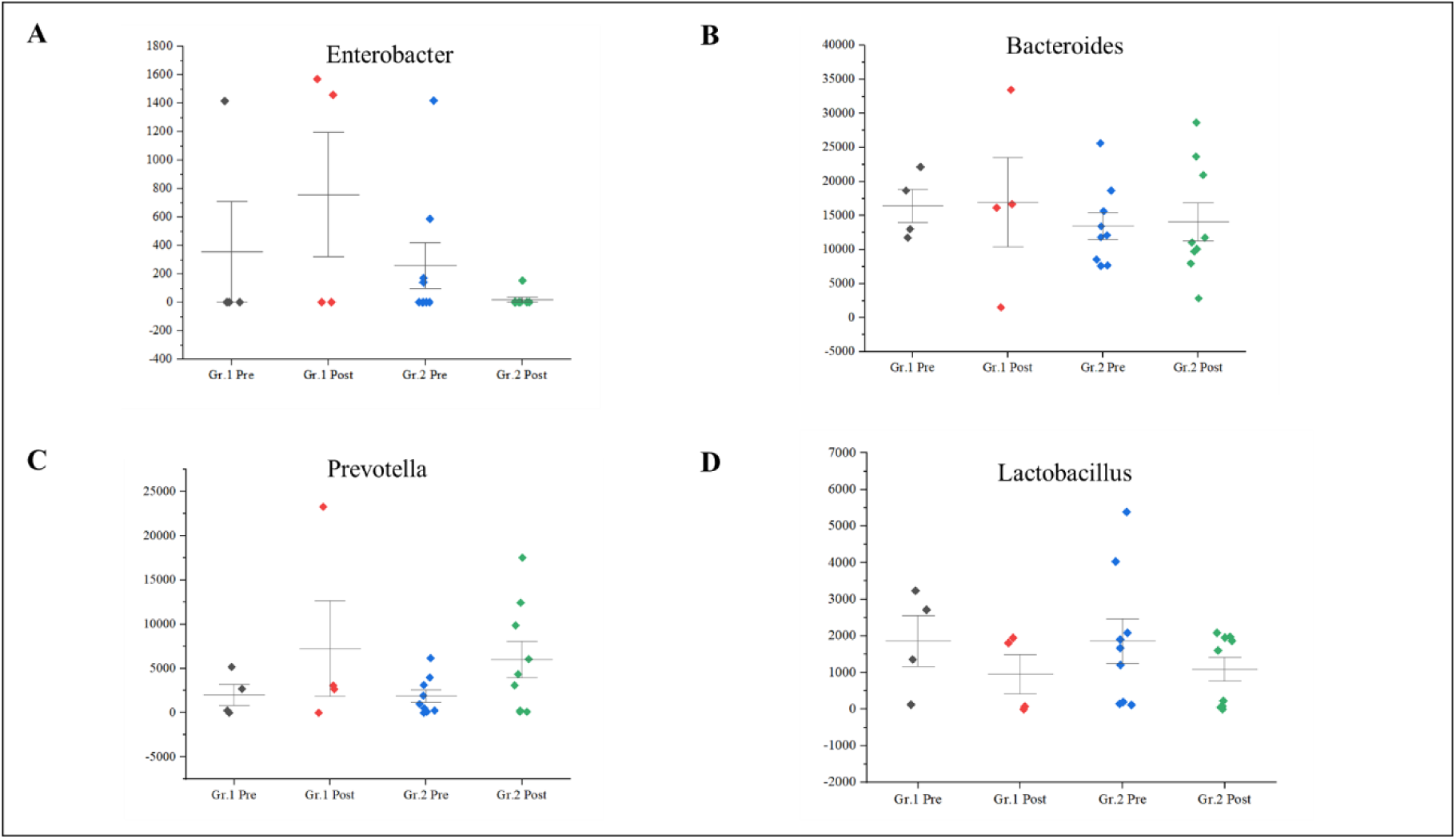
(A) Decrease in abundance of Enterobacteria in Group 2 compared to Group (Gr.) 1, postintervention. (B) Decrease in abundance of Bacteroides in Group (Gr.) 2, which had increased in Group 1 postintervention. (C) Increase in Prevotella in Group 1 and Group 2 postintervention and (D) Decrease in lactobacillus in Group 1 and Group 2 postintervention.

**Figure 4:**
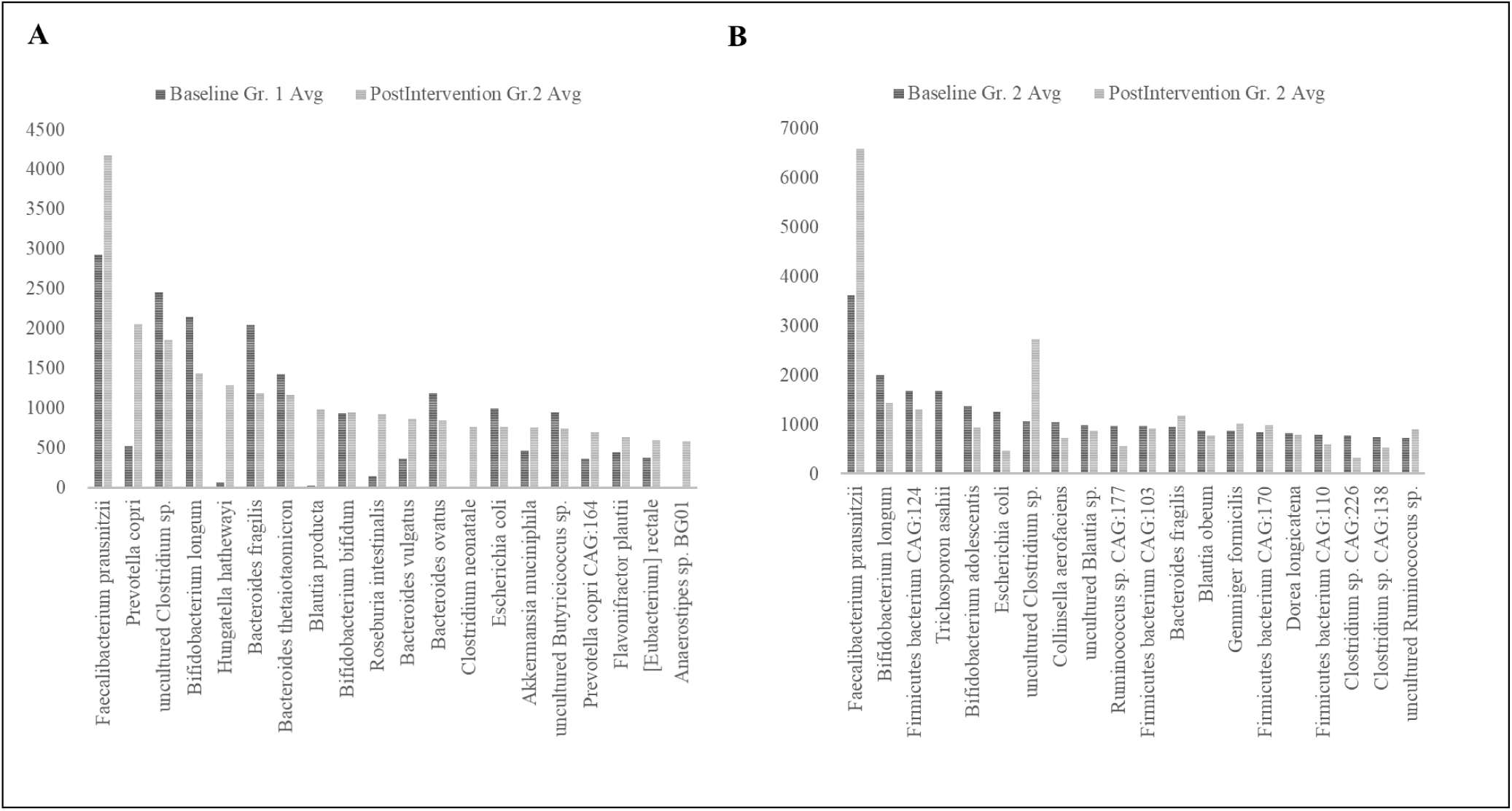
Species Abundance of major species analysed; (A) Group (Gr.) 1 at baseline versus postintervention and (B) Group (Gr.) 2 at baseline versus postintervention.

**Figure 5:**
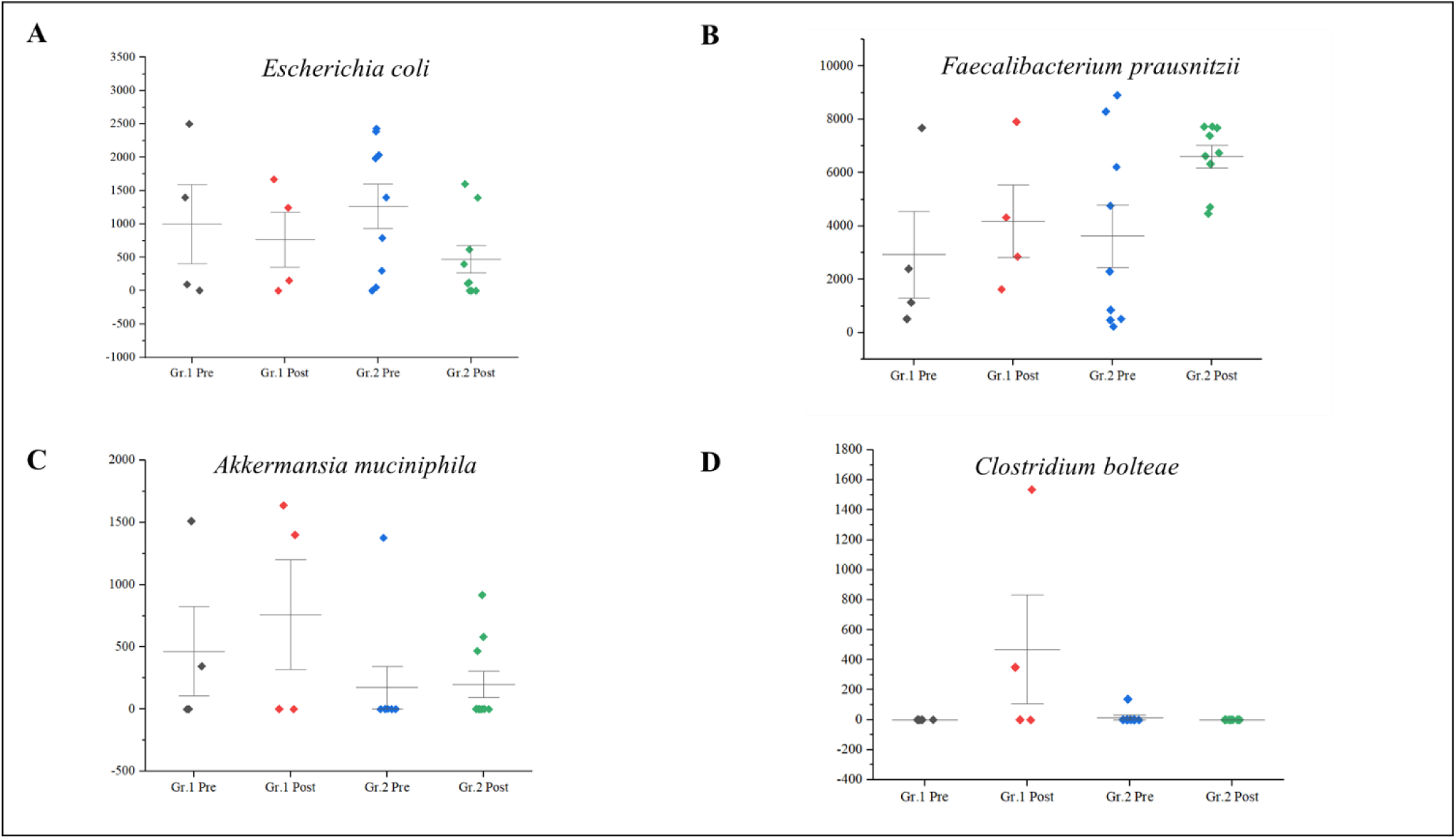
(A) Decrease in abundance of E. coli was significant in Group (Gr.) 2 compared to Group (Gr.) 1, postintervention. (B) Significant increase in abundance of Faecalibacterium prausnitzii in Group 2 compared to Group 1 postintervention. (C) Increase in Akkermansia muciniphila in Group 1 but decrease in Group 2 postintervention. (D) Increase in Clostridium bolteae CAG:59 in Group 1 but decrease in Group 2 postintervention.

The data of the genus and species level abundance in Gr. 1 and Gr. 2 are presented in supplementary tables 1-4.

## Discussion

In this study, following earlier reports of clinical improvement in terms of sleep [11], behavioural pattern [12], plasma αSyn [12], and serum melatonin increase [11], gut dysbiosis has been shown to have a strong correlation with the severity of symptoms in ASD [13]. We evaluated and compared the gut microbiota of the subjects who were supplemented with AFO-202-derived 1,3-1,6 beta glucan with those who did not take the supplement. Several studies have reported the differences in the gut microbiota between children with ASD and neurotypical children. Reduced number of bifidobacterial and increased Clostridium spp., Desulfovibrio spp., Sutterella spp., and/or Veillonellacea was reported by Souza et al [14]. Tomova et al [15] reported a change in Bacteroidetes/Firmicutes ratio and an increase in bifidobacterial numbers after probiotic administration. An exclusion diet and a 6-week prebiotic intervention demonstrated lower abundance of Bifidobacterium spp. and Veillonellaceae family and higher abundance of Faecalibacterium prausnitzii and Bacteroides spp. [13]. Faecalibacterium, Ruminococcus, and Bifidobacterium were relatively less abundant, whereas Caloramator, Sarcina, Sutterella ceae, and Enterobacteriaceae were more abundant in children with ASD[16]. In addition, lower abundances of the genera Prevotella, Coprococcus, and unclassified Veillonellaceae have been reported [17]. Among these bacteria, increased Bacteroidaceae, Prevotellaceae, and Ruminococcaceae and decreased Prevotella copri, Faecalibacterium prausnitzii, and Haemophilus parainfluenzae have been reported [18,19]. In the present study, in line with these reports, the shift of the gut microbiome was towards a beneficial spectrum in Group 2 (Nichi Glucan) because there was a decrease in Enterobacter, lactobacillus, Escherichia coli, Akkermansia muciniphila CAG:154, Blautia spp., Coprobacillus sp., several clostridium spp., and Clostridium bolteae CAG:59, with an increase in the abundance of Bacteroides, Prevotella, Faecalibacterium prausnitzii, and Prevotella copri. Desulfovibrio Bacteria which have been reported to be associated with PD [20] decreased in the Gr. 2.

In particular, Enterobacteria and E. coli significantly decreased in Group 2 compared to Group 1 after the intervention. Gram-negative enteric bacteria such as the Enterobacter and E coli secrete the amyloid curli that constitutes 85% of the extracellular matrix of enteric biofilms. The curli has similarities and associations with pathological and immunomodulatory human amyloids such as amyloid-β implicated in AD, αSyn involved in ASD and PD, and serum amyloid A associated with neuroinflammation [21]. Curli causes misfolding [22] and accumulation of the neuronal protein αSyn in the form of insoluble amyloid aggregations, leading to inflammation and neuronal dysfunction that is central to pathogenesis of Lewy-body-associated synucleinopathies, including PD and AD. Curli-producing bacteria also increase the production and aggregation of the amyloid protein αSyn, which has been shown to propagate in a prion-like fashion from the gut to the brain via the vagus nerve and/or spinal cord, thus culminating in the neurological disorders such as ASD [22]. In this study, the significant decrease in Enterobacter and E-coli will thus be of benefit in these synculeopathies. Before the start of this study, the objective to study αSyn was to understand the effects of the beta glucan supplementation on Synaptic imbalance in presynaptic terminals, observed in ASD. The study results showed that plasma levels of αSyn increased in Group 2 compared with Group 1 along with improvement in Childhood Autism Rating Scale score and sleep pattern which to our knowledge is the first of its kind intervention producing an observable change in the plasma synuclein levels [22]. The study of the gut microbiome has offered further new insights wherein Enterobacter increasing curli protein and αSyn deposition in the enteric nervous system having been controlled by the beta glucan food supplement, the increase in plasma αSyn levels point out to the disintegration of the amyloid deposits leading to these αSyn entering the blood stream. Indeed, natural killer (NK) cells have been shown to act as efficient scavengers of abnormal α-Syn aggregates [23], and the AFO-202 beta glucan has a proven capability to increase and activate NK cells [24], which could be another probable mechanism contributing to the increased αSyn levels in the plasma, apart from positive clinical outcomes in these children with ASD. This result highlights NK cell’s potential as a promising therapeutic strategy for prophylaxis and prevention of brain disorders, and they are likely to be used for such αSyn accumulation and propagation, after relevant research on their specific pathways and variations in their capability. The NK cells have been proven to clear the amyloid deposits peripherally though not macrophages [25]. In the central nervous system, such a role is played by the microglia [26]. βeta-glucans also rejuvenate microglia [27] that have been shown to scavenge amyloid deposits in the brain and CNS [28], thus proving to be a wholesome therapeutic strategy for neurodevelopmental and neurodegenerative diseases. Altered α-Syn protein misfolding spreading to anatomically connected regions in a prion-like manner and mediating neurodegenerative diseases such as PD [29] and the increased risk of children with ASD in developing PD at a later stage [30] suggests further research into these converging pathogenic pathways of neurodevelopmental and neurodegenerative diseases is needed as well as suggests that this safety-proven food supplement is a preventive strategy in subjects with ASD against PD. Other than research into normal and abnormal α-syn being warranted, studying the implications of soluble and insoluble α-syn [31] is important because the proportion of insoluble α-syn that was phosphorylated at Ser129 was reported to be significantly higher in brain tissue from PD patients. In addition, cell lines such as the SH-SYHY neuroblastoma [31] will reveal the correlation of the various α-syn with the severity of symptoms and pathogenesis in these neurodegenerative diseases, apart from helping to develop novel disease-modifying strategies employing such simple nutritional supplementation. The microbiota reconstituted in a beneficial manner in the present study with AFO-202 beta glucan must be further progressed into research to study the effects of other variants of A. pullulans beta glucan that have been shown to be anti-inflammatory. Such research could lead to mechanistic insight into the molecular pathways from the local immune responses in the gut leading to systemic inflammation and, eventually, to organ-specific autoimmunity of the CS in neuro-inflammatory conditions such as MS.

## Conclusion

Favourable reconstitution of the gut microbiota after consumption of AFO-202 beta glucan in children with ASD has been demonstrated in this study, apart from the clinical improvement already reported. The decrease in Enterobacteria demonstrates the potential of this beta-glucan supplementation for neurodevelopmental conditions such as ASD as well as neurodegenerative disorders such as PD and AD, with converging pathways of amyloid accumulations and propagation, warranting larger clinical studies and research to recommend this as a routine food supplement or an adjunct to existing therapies for prevention and management of both neurodevelopmental and neurodegenerative diseases.

## Data Availability

All data produced in the present work are contained in the manuscript

## Acknowledgements

The authors thank

1. The Government of Japan and the Prefectural Government of Yamanashi for a special loan and M/s Yamanashi Chuo Bank for processing the transactions.
2. Mr. Mohan Ponnusamy, Dr. Madhankumar & staff of Kenmax for their assistance during the clinical study and data collection of the manuscript.
3. Mr. Takashi Onaka, Mr. Yasunori Ikeue, Mr. Mitsuru Nagataki (Sophy Inc, Kochi, Japan), for necessary technical clarifications.
4. Mr. Yoshio Morozumi and Ms. Yoshiko Amikura of GN Corporation, Japan for their liaison assistance with the conduct of the study.
5. Loyola-ICAM College of Engineering and Technology (LICET) for their support to our research work.

