## Supplementary Table for "Beneficial reconstitution of gut microbiota and control of alpha-synuclein and curli-amyloids-producing enterobacteria, by beta 1,3-1,6 glucans in a clinical pilot study of autism and potentials in neurodegenerative diseases"

**Table 1: Genus level: Mean and percentage abundance at baseline and post-intervention in Gr.1 (Control)**

| **S.No** | **Genus** | **Mean abundance** | | **Percentage of abundance** | |
| --- | --- | --- | --- | --- | --- |
|  |  | **Baseline** | **Post-Intervention** | **Baseline** | **Post-Intervention** |
| 1 | Bacteroides | 16395 | 16956 | 16.84 | 19.09 |
| 2 | Prevotella | 2019 | 7252 | 2.07 | 8.16 |
| 3 | Clostridium | 6277 | 5792 | 6.45 | 6.52 |
| 4 | Faecalibacterium | 4646 | 5398 | 4.77 | 6.08 |
| 5 | Bifidobacterium | 7375 | 4809 | 7.58 | 5.41 |
| 6 | Blautia | 6123 | 4175 | 6.29 | 4.70 |
| 7 | Roseburia | 3257 | 3564 | 3.35 | 4.01 |
| 8 | Eubacterium | 2268 | 3016 | 2.33 | 3.40 |
| 9 | Ruminococcus | 3484 | 2066 | 3.58 | 2.33 |
| 10 | Lachnoclostridium | 1289 | 1682 | 1.32 | 1.89 |
| 11 | Hungatella | 66 | 1381 | 0.07 | 1.55 |
| 12 | Dialister | 983 | 1380 | 1.01 | 1.55 |
| 13 | Klebsiella | 1121 | 1357 | 1.15 | 1.53 |
| 14 | Alistipes | 2487 | 1352 | 2.55 | 1.52 |
| 15 | Enterococcus | 4721 | 1270 | 4.85 | 1.43 |
| 16 | Veillonella | 0 | 1244 | 0.00 | 1.40 |
| 17 | Akkermansia | 827 | 1126 | 0.85 | 1.27 |
| 18 | Streptococcus | 384 | 1093 | 0.39 | 1.23 |
| 19 | Megasphaera | 64 | 1084 | 0.07 | 1.22 |
| 20 | Anaerostipes | 393 | 1020 | 0.40 | 1.15 |
| 21 | Parabacteroides | 1686 | 971 | 1.73 | 1.09 |
| 22 | Lactobacillus | 1857 | 956 | 1.91 | 1.08 |
| 23 | Coprococcus | 897 | 915 | 0.92 | 1.03 |
| 24 | Dorea | 1352 | 862 | 1.39 | 0.97 |
| 25 | Butyricicoccus | 1339 | 835 | 1.38 | 0.94 |
| 26 | Flavonifractor | 669 | 825 | 0.69 | 0.93 |
| 27 | Escherichia | 1062 | 822 | 1.09 | 0.92 |
| 28 | Odoribacter | 1036 | 803 | 1.06 | 0.90 |
| 29 | Enterobacter | **355** | **757** | **0.36** | **0.85** |
| 30 | Subdoligranulum | 802 | 597 | 0.82 | 0.67 |
| 31 | Oscillibacter | 699 | 569 | 0.72 | 0.64 |
| 32 | Bilophila | 0 | 552 | 0.00 | 0.62 |
| 33 | Eggerthella | 829 | 548 | 0.85 | 0.62 |
| 34 | Collinsella | 1617 | 541 | 1.66 | 0.61 |
| 35 | Turicibacter | 0 | 415 | 0.00 | 0.47 |
| 36 | Mitsuokella | 0 | 409 | 0.00 | 0.46 |
| 37 | Catenibacterium | 540 | 403 | 0.55 | 0.45 |
| 38 | Fusicatenibacter | 434 | 392 | 0.45 | 0.44 |
| 39 | Romboutsia | 206 | 357 | 0.21 | 0.40 |
| 40 | Haemophilus | 88 | 346 | 0.09 | 0.39 |
| 41 | Gemmiger | 524 | 343 | 0.54 | 0.39 |
| 42 | Sutterella | 139 | 301 | 0.14 | 0.34 |
| 43 | Lachnospira | 0 | 294 | 0.00 | 0.33 |
| 44 | Mycoplasma | 292 | 251 | 0.30 | 0.28 |
| 45 | Holdemanella | 585 | 199 | 0.60 | 0.22 |
| 46 | Parasutterella | 0 | 197 | 0.00 | 0.22 |
| 47 | Weissella | 173 | 177 | 0.18 | 0.20 |
| 48 | Ruminiclostridium | 174 | 148 | 0.18 | 0.17 |
| 49 | Chlamydia | 133 | 147 | 0.14 | 0.16 |
| 50 | Dakarella | 97 | 147 | 0.10 | 0.16 |
| 51 | Lactococcus | 144 | 144 | 0.15 | 0.16 |
| 52 | Clostridioides | 107 | 126 | 0.11 | 0.14 |
| 53 | Terrisporobacter | 0 | 123 | 0.00 | 0.14 |
| 54 | Coprobacillus | 224 | 114 | 0.23 | 0.13 |
| 55 | Adlercreutzia | 405 | 101 | 0.42 | 0.11 |
| 56 | Butyrivibrio | 121 | 90 | 0.12 | 0.10 |
| 57 | Anaerotruncus | 135 | 81 | 0.14 | 0.09 |
| 58 | Bacillus | 262 | 69 | 0.27 | 0.08 |
| 59 | Agathobaculum | 54 | 67 | 0.05 | 0.08 |
| 60 | Shigella | 53 | 60 | 0.05 | 0.07 |
| 61 | Tyzzerella | 13 | 59 | 0.01 | 0.07 |
| 62 | Oribacterium | 145 | 52 | 0.15 | 0.06 |
| 63 | Intestinimonas | 91 | 50 | 0.09 | 0.06 |
| 64 | Citrobacter | 0 | 49 | 0.00 | 0.06 |
| 65 | Fusobacterium | 0 | 46 | 0.00 | 0.05 |
| 66 | Acholeplasma | 0 | 41 | 0.00 | 0.05 |
| 67 | Pseudoflavonifractor | 68 | 40 | 0.07 | 0.05 |
| 68 | Raoultella | 0 | 32 | 0.00 | 0.04 |
| 69 | Muribaculum | 0 | 31 | 0.00 | 0.03 |
| 70 | Acidiphilium | 27 | 29 | 0.03 | 0.03 |
| 71 | Paenibacillus | 110 | 28 | 0.11 | 0.03 |
| 72 | Acidaminococcus | 0 | 25 | 0.00 | 0.03 |
| 73 | Salmonella | 0 | 22 | 0.00 | 0.03 |
| 74 | Anaeromassilibacillus | 179 | 22 | 0.18 | 0.02 |
| 75 | Pantoea | 0 | 21 | 0.00 | 0.02 |
| 76 | Intestinibacter | 0 | 21 | 0.00 | 0.02 |
| 77 | Eisenbergiella | 85 | 20 | 0.09 | 0.02 |
| 78 | Prevotellamassilia | 0 | 20 | 0.00 | 0.02 |
| 79 | Anaerotignum | 0 | 17 | 0.00 | 0.02 |
| 80 | Selenomonas | 0 | 16 | 0.00 | 0.02 |
| 81 | Actinomyces | 0 | 16 | 0.00 | 0.02 |
| 82 | Paraprevotella | 26 | 10 | 0.03 | 0.01 |
| 83 | Porphyromonas | 0 | 9 | 0.00 | 0.01 |
| 84 | Barnesiella | 129 | 0 | 0.13 | 0.00 |
| 85 | Anaerococcus | 141 | 0 | 0.14 | 0.00 |
| 86 | Stenotrophomonas | 13 | 0 | 0.01 | 0.00 |
| 87 | Finegoldia | 82 | 0 | 0.08 | 0.00 |
| 88 | Pediococcus | 10 | 0 | 0.01 | 0.00 |
| 89 | Solobacterium | 27 | 0 | 0.03 | 0.00 |
| 90 | Peptoniphilus | 929 | 0 | 0.95 | 0.00 |
| 91 | Urinacoccus | 144 | 0 | 0.15 | 0.00 |
| 92 | Enterorhabdus | 46 | 0 | 0.05 | 0.00 |
| 93 | Slackia | 69 | 0 | 0.07 | 0.00 |
| 94 | Achromobacter | 59 | 0 | 0.06 | 0.00 |
| 95 | Lysinibacillus | 1130 | 0 | 1.16 | 0.00 |
| 96 | Olsenella | 192 | 0 | 0.20 | 0.00 |
| 97 | Comamonas | 131 | 0 | 0.13 | 0.00 |
| 98 | Ruthenibacterium | 63 | 0 | 0.06 | 0.00 |
| 99 | Viridibacillus | 40 | 0 | 0.04 | 0.00 |
| 100 | Rummeliibacillus | 603 | 0 | 0.62 | 0.00 |
| 101 | Gordonibacter | 735 | 0 | 0.75 | 0.00 |
| 102 | Drancourtella | 19 | 0 | 0.02 | 0.00 |
| 103 | Monoglobus | 24 | 0 | 0.02 | 0.00 |
| 104 | Lagierella | 39 | 0 | 0.04 | 0.00 |
| 105 | Peptococcus | 27 | 0 | 0.03 | 0.00 |
| 106 | Senegalimassilia | 122 | 0 | 0.13 | 0.00 |
| 107 | Libanicoccus | 25 | 0 | 0.03 | 0.00 |
| 108 | Urmitella | 24 | 0 | 0.02 | 0.00 |
| 109 | Paraclostridium | 19 | 0 | 0.02 | 0.00 |
| 110 | Acinetobacter | 1273 | 0 | 1.31 | 0.00 |
| 111 | Anaerocolumna | 9 | 0 | 0.01 | 0.00 |
| 112 | Marvinbryantia | 22 | 0 | 0.02 | 0.00 |
| 113 | Robinsoniella | 50 | 0 | 0.05 | 0.00 |
| 114 | Neglecta | 33 | 0 | 0.03 | 0.00 |
| 115 | Butyricimonas | 180 | 0 | 0.19 | 0.00 |
| 116 | unknown | 6947 | 5101 | 7.14 | 5.74 |

**Table 2: Genus level: Mean and percentage abundance at baseline and post-intervention in Gr.2 (Nichi Glucan)**

| **S.No** | **Genus** | **Mean abundance** | | **Percentage of abundance** | |
| --- | --- | --- | --- | --- | --- |
|  |  | **Baseline** | **Post-Intervention** | **Baseline** | **Post-Intervention** |
| 1 | **Bacteroides** | 13463 | 14088 | 11.60 | 11.43 |
| 2 | **Clostridium** | 8728 | 12105 | 7.52 | 9.82 |
| 3 | **Bifidobacterium** | 6771 | 5625 | 5.84 | 4.57 |
| 4 | **Faecalibacterium** | 5291 | 9278 | 4.56 | 7.53 |
| 5 | **Enterococcus** | 4583 | 40 | 3.95 | 0.03 |
| 6 | **Eubacterium** | 4380 | 5007 | 3.78 | 4.06 |
| 7 | **Ruminococcus** | 4265 | 6143 | 3.68 | 4.99 |
| 8 | **Blautia** | 4196 | 3973 | 3.62 | 3.22 |
| 9 | **Alistipes** | 3187 | 2268 | 2.75 | 1.84 |
| 10 | **Collinsella** | 2542 | 1590 | 2.19 | 1.29 |
| 11 | **Parabacteroides** | 2419 | 1307 | 2.08 | 1.06 |
| 12 | **Oscillibacter** | 1956 | 2542 | 1.69 | 2.06 |
| 13 | **Prevotella** | 1887 | 5979 | 1.63 | 4.85 |
| 14 | **Lactobacillus** | 1860 | 1093 | 1.60 | 0.89 |
| 15 | **Roseburia** | 1756 | 7932 | 1.51 | 6.44 |
| 16 | **Trichosporon** | 1683 | 0 | 1.45 | 0.00 |
| 17 | **Dorea** | 1592 | 1585 | 1.37 | 1.29 |
| 18 | **Lachnoclostridium** | 1360 | 1056 | 1.17 | 0.86 |
| 19 | **Olsenella** | 1354 | 246 | 1.17 | 0.20 |
| 20 | **Escherichia** | 1346 | 505 | 1.16 | 0.41 |
| 21 | **Klebsiella** | 1067 | 80 | 0.92 | 0.06 |
| 22 | **Gemmiger** | 970 | 1161 | 0.84 | 0.94 |
| 23 | **Streptococcus** | 925 | 521 | 0.80 | 0.42 |
| 24 | **Coprococcus** | 910 | 1228 | 0.78 | 1.00 |
| 25 | **Subdoligranulum** | 904 | 1112 | 0.78 | 0.90 |
| 26 | **Eggerthella** | 871 | 190 | 0.75 | 0.15 |
| 27 | **Dialister** | 765 | 1062 | 0.66 | 0.86 |
| 28 | **Hungatella** | 732 | 175 | 0.63 | 0.14 |
| 29 | **Lysinibacillus** | 709 | 0 | 0.61 | 0.00 |
| 30 | **Acinetobacter** | 677 | 0 | 0.58 | 0.00 |
| 31 | **Pediococcus** | 662 | 0 | 0.57 | 0.00 |
| 32 | **Holdemanella** | 649 | 546 | 0.56 | 0.44 |
| 33 | **Butyricicoccus** | 623 | 1257 | 0.54 | 1.02 |
| 34 | **Catenibacterium** | 620 | 470 | 0.53 | 0.38 |
| 35 | **Cloacibacillus** | 602 | 265 | 0.52 | 0.21 |
| 36 | **Pichia** | 588 | 0 | 0.51 | 0.00 |
| 37 | **Odoribacter** | 577 | 271 | 0.50 | 0.22 |
| 38 | **Rummeliibacillus** | 555 | 0 | 0.48 | 0.00 |
| 39 | **Weissella** | 555 | 24 | 0.48 | 0.02 |
| 40 | **Flavonifractor** | 545 | 729 | 0.47 | 0.59 |
| 41 | **Senegalimassilia** | 505 | 56 | 0.43 | 0.05 |
| 42 | **Megasphaera** | 464 | 1203 | 0.40 | 0.98 |
| 43 | **Romboutsia** | 463 | 130 | 0.40 | 0.11 |
| 44 | **Desulfovibrio** | 461 | 344 | 0.40 | 0.28 |
| 45 | **Fusicatenibacter** | 452 | 641 | 0.39 | 0.52 |
| 46 | **Sutterella** | 436 | 780 | 0.38 | 0.63 |
| 47 | **Mycoplasma** | 424 | 265 | 0.37 | 0.21 |
| 48 | **Peptoniphilus** | 413 | 0 | 0.36 | 0.00 |
| 49 | **Ruminiclostridium** | 411 | 387 | 0.35 | 0.31 |
| 50 | **Anaerotruncus** | 410 | 485 | 0.35 | 0.39 |
| 51 | **Butyrivibrio** | 404 | 315 | 0.35 | 0.26 |
| 52 | **Methanobrevibacter** | 395 | 368 | 0.34 | 0.30 |
| 53 | **Megamonas** | 392 | 501 | 0.34 | 0.41 |
| 54 | **Bacillus** | 326 | 150 | 0.28 | 0.12 |
| 55 | **Butyricimonas** | 319 | 39 | 0.28 | 0.03 |
| 56 | **Anaerostipes** | 318 | 485 | 0.27 | 0.39 |
| 57 | **Phascolarctobacterium** | 317 | 464 | 0.27 | 0.38 |
| 58 | **Akkermansia** | 305 | 498 | 0.26 | 0.40 |
| 59 | **Gordonibacter** | 304 | 0 | 0.26 | 0.00 |
| 60 | **Coprobacillus** | 295 | 438 | 0.25 | 0.36 |
| 61 | **Coraliomargarita** | 294 | 339 | 0.25 | 0.28 |
| 62 | **Methanosphaera** | 282 | 0 | 0.24 | 0.00 |
| 63 | **Enterobacter** | **257** | **17** | **0.22** | **0.01** |
| 64 | **Pyramidobacter** | 231 | 15 | 0.20 | 0.01 |
| 65 | **Acidaminococcus** | 230 | 269 | 0.20 | 0.22 |
| 66 | **Paenibacillus** | 229 | 184 | 0.20 | 0.15 |
| 67 | **Acidiphilium** | 219 | 555 | 0.19 | 0.45 |
| 68 | **Adlercreutzia** | 210 | 334 | 0.18 | 0.27 |
| 69 | **Actinomyces** | 191 | 4 | 0.16 | 0.00 |
| 70 | **Succinatimonas** | 190 | 383 | 0.16 | 0.31 |
| 71 | **Libanicoccus** | 188 | 119 | 0.16 | 0.10 |
| 72 | **Angelakisella** | 184 | 79 | 0.16 | 0.06 |
| 73 | **Pseudoflavonifractor** | 183 | 173 | 0.16 | 0.14 |
| 74 | **Chlamydia** | 176 | 130 | 0.15 | 0.11 |
| 75 | **Anaeromassilibacillus** | 175 | 135 | 0.15 | 0.11 |
| 76 | **Intestinimonas** | 157 | 174 | 0.14 | 0.14 |
| 77 | **Duodenibacillus** | 150 | 181 | 0.13 | 0.15 |
| 78 | **Oribacterium** | 145 | 103 | 0.13 | 0.08 |
| 79 | **Clostridioides** | 129 | 64 | 0.11 | 0.05 |
| 80 | **Barnesiella** | 118 | 84 | 0.10 | 0.07 |
| 81 | **Slackia** | 109 | 0 | 0.09 | 0.00 |
| 82 | **Eisenbergiella** | 95 | 112 | 0.08 | 0.09 |
| 83 | **Acidovorax** | 93 | 0 | 0.08 | 0.00 |
| 84 | **Lactococcus** | 92 | 0 | 0.08 | 0.00 |
| 85 | **Paraprevotella** | 91 | 16 | 0.08 | 0.01 |
| 86 | **Shigella** | 90 | 90 | 0.08 | 0.07 |
| 87 | **Oxalobacter** | 87 | 0 | 0.08 | 0.00 |
| 88 | **Selenomonas** | 84 | 231 | 0.07 | 0.19 |
| 89 | **Turicibacter** | 74 | 36 | 0.06 | 0.03 |
| 90 | **Haemophilus** | 74 | 196 | 0.06 | 0.16 |
| 91 | **Pygmaiobacter** | 69 | 0 | 0.06 | 0.00 |
| 92 | **Urinacoccus** | 64 | 0 | 0.06 | 0.00 |
| 93 | **Bilophila** | 63 | 613 | 0.05 | 0.50 |
| 94 | **Allisonella** | 62 | 54 | 0.05 | 0.04 |
| 95 | **Ruthenibacterium** | 51 | 69 | 0.04 | 0.06 |
| 96 | **Raoultibacter** | 50 | 0 | 0.04 | 0.00 |
| 97 | **Comamonas** | 44 | 0 | 0.04 | 0.00 |
| 98 | **Neglecta** | 42 | 37 | 0.04 | 0.03 |
| 99 | **Atopobium** | 40 | 11 | 0.03 | 0.01 |
| 100 | **Christensenella** | 38 | 41 | 0.03 | 0.03 |
| 101 | **Massilimaliae** | 37 | 32 | 0.03 | 0.03 |
| 102 | **Finegoldia** | 36 | 0 | 0.03 | 0.00 |
| 103 | **Treponema** | 36 | 48 | 0.03 | 0.04 |
| 104 | **Robinsoniella** | 36 | 0 | 0.03 | 0.00 |
| 105 | **Enterorhabdus** | 36 | 40 | 0.03 | 0.03 |
| 106 | **Viridibacillus** | 36 | 0 | 0.03 | 0.00 |
| 107 | **Peptococcus** | 35 | 0 | 0.03 | 0.00 |
| 108 | **Fusobacterium** | 34 | 30 | 0.03 | 0.02 |
| 109 | **Terrisporobacter** | 34 | 7 | 0.03 | 0.01 |
| 110 | **Tyzzerella** | 33 | 171 | 0.03 | 0.14 |
| 111 | **Erysipelatoclostridium** | 32 | 0 | 0.03 | 0.00 |
| 112 | **Sporobacter** | 32 | 36 | 0.03 | 0.03 |
| 113 | **Mitsuokella** | 31 | 432 | 0.03 | 0.35 |
| 114 | **Cutaneotrichosporon** | 31 | 0 | 0.03 | 0.00 |
| 115 | **Marvinbryantia** | 31 | 28 | 0.03 | 0.02 |
| 116 | **Corallococcus** | 30 | 143 | 0.03 | 0.12 |
| 117 | **Anaerotignum** | 28 | 18 | 0.02 | 0.01 |
| 118 | **Prevotellamassilia** | 24 | 158 | 0.02 | 0.13 |
| 119 | **Anaerofilum** | 24 | 35 | 0.02 | 0.03 |
| 120 | **Synergistes** | 22 | 0 | 0.02 | 0.00 |
| 121 | **Alloprevotella** | 22 | 22 | 0.02 | 0.02 |
| 122 | **Isoptericola** | 22 | 0 | 0.02 | 0.00 |
| 123 | **Anaerococcus** | 21 | 0 | 0.02 | 0.00 |
| 124 | **Propionibacterium** | 20 | 0 | 0.02 | 0.00 |
| 125 | **Leuconostoc** | 20 | 0 | 0.02 | 0.00 |
| 126 | **Brachyspira** | 19 | 95 | 0.02 | 0.08 |
| 127 | **Lachnospira** | 18 | 386 | 0.02 | 0.31 |
| 128 | **Lagierella** | 17 | 0 | 0.02 | 0.00 |
| 129 | **Mogibacterium** | 16 | 0 | 0.01 | 0.00 |
| 130 | **Agathobaculum** | 15 | 61 | 0.01 | 0.05 |
| 131 | **Pseudobutyrivibrio** | 15 | 35 | 0.01 | 0.03 |
| 132 | **Hydrogenoanaerobacterium** | 14 | 14 | 0.01 | 0.01 |
| 133 | **Holdemania** | 14 | 0 | 0.01 | 0.00 |
| 134 | **Desulfotomaculum** | 14 | 0 | 0.01 | 0.00 |
| 135 | **Veillonella** | 14 | 324 | 0.01 | 0.26 |
| 136 | **Succinivibrio** | 14 | 32 | 0.01 | 0.03 |
| 137 | **Anaerocolumna** | 14 | 14 | 0.01 | 0.01 |
| 138 | **Mobilibacterium** | 12 | 0 | 0.01 | 0.00 |
| 139 | **Arabia** | 12 | 0 | 0.01 | 0.00 |
| 140 | **Enorma** | 11 | 0 | 0.01 | 0.00 |
| 141 | **Candida <Debaryomycetaceae>** | 11 | 0 | 0.01 | 0.00 |
| 142 | **Intestinibacter** | 11 | 13 | 0.01 | 0.01 |
| 143 | **Kurthia** | 10 | 0 | 0.01 | 0.00 |
| 144 | **Paeniclostridium** | 9 | 0 | 0.01 | 0.00 |
| 145 | **Paraclostridium** | 8 | 0 | 0.01 | 0.00 |
| 146 | **Listeria** | 8 | 0 | 0.01 | 0.00 |
| 147 | **Kwoniella** | 8 | 0 | 0.01 | 0.00 |
| 148 | **Cryptococcus** | 7 | 0 | 0.01 | 0.00 |
| 149 | **Fournierella** | 0 | 23 | 0.00 | 0.02 |
| 150 | **Sellimonas** | 0 | 23 | 0.00 | 0.02 |
| 151 | **Blastocystis** | 0 | 34 | 0.00 | 0.03 |
| 152 | **Acholeplasma** | 0 | 126 | 0.00 | 0.10 |
| 153 | **Azospirillum** | 0 | 162 | 0.00 | 0.13 |
| 154 | **Porphyromonas** | 0 | 7 | 0.00 | 0.01 |
| 155 | **Emergencia** | 0 | 14 | 0.00 | 0.01 |
| 156 | **Acetobacter** | 0 | 13 | 0.00 | 0.01 |
| 157 | **Dakarella** | 0 | 64 | 0.00 | 0.05 |
| 158 | **Parasutterella** | 0 | 133 | 0.00 | 0.11 |
| 159 | **Elusimicrobium** | 0 | 128 | 0.00 | 0.10 |
| 160 | **Drancourtella** | 0 | 32 | 0.00 | 0.03 |
| 161 | **Anaerovorax** | 0 | 15 | 0.00 | 0.01 |
| 162 | **Caldicoprobacter** | 0 | 15 | 0.00 | 0.01 |
| 163 | **unknown** | 12707 | 16408 | 10.95 | 13.32 |

**Table 43: Species level: Mean abundance at baseline and post-intervention in**

**Gr.1 (Control)**

| **S.No** | **Species** | **Mean abundance** | |
| --- | --- | --- | --- |
|  |  | **Baseline** | **Post-Intervention** |
| 1 | **Prevotella copri** | 530 | 2058 |
| 2 | **uncultured Clostridium sp.** | 2459 | 1860 |
| 3 | **Bifidobacterium longum** | 2150 | 1433 |
| 4 | **Hungatella hathewayi** | 65 | 1287 |
| 5 | **Bacteroides fragilis** | 2052 | 1189 |
| 6 | **Bacteroides thetaiotaomicron** | 1432 | 1172 |
| 7 | **Blautia producta** | 25 | 985 |
| 8 | **Bifidobacterium bifidum** | 941 | 946 |
| 9 | **Roseburia intestinalis** | 144 | 930 |
| 10 | **Bacteroides vulgatus** | 364 | 866 |
| 11 | **Bacteroides ovatus** | 1186 | 846 |
| 12 | **Clostridium neonatale** | 0 | 770 |
| 13 | **Escherichia coli** | 998 | 767 |
| 14 | **Akkermansia muciniphila** | 464 | 760 |
| 15 | **uncultured Butyricicoccus sp.** | 948 | 751 |
| 16 | **Prevotella copri CAG:164** | 362 | 700 |
| 17 | **Flavonifractor plautii** | 445 | 640 |
| 18 | **[Eubacterium] rectale** | 380 | 594 |
| 19 | **Anaerostipes sp. BG01** | 0 | 584 |
| 20 | **Dialister sp. CAG:357** | 0 | 540 |
| 21 | **Lactobacillus ruminis** | 628 | 536 |
| 22 | **Bacteroides uniformis** | 265 | 519 |
| 23 | **Roseburia faecis** | 625 | 507 |
| 24 | **Prevotella sp. CAG:386** | 61 | 481 |
| 25 | **Eubacterium sp. CAG:252** | 0 | 472 |
| 26 | **[Clostridium] bolteae** | 0 | 471 |
| 27 | **[Eubacterium] eligens** | 0 | 470 |
| 28 | **Bifidobacterium adolescentis** | 574 | 466 |
| 29 | **Klebsiella pneumoniae** | 397 | 461 |
| 30 | **Prevotella sp. CAG:1092** | 0 | 457 |
| 31 | **Dialister succinatiphilus** | 412 | 454 |
| 32 | **Clostridium sp. AT4** | 12 | 450 |
| 33 | **Prevotella sp. CAG:604** | 154 | 445 |
| 34 | **Firmicutes bacterium CAG:124** | 806 | 439 |
| 35 | **uncultured Blautia sp.** | 1136 | 435 |
| 36 | **Bacteroides xylanisolvens** | 402 | 414 |
| 37 | **Bilophila wadsworthia** | 0 | 412 |
| 38 | **Blautia obeum** | 1024 | 410 |
| 39 | **Prevotella sp. 885** | 48 | 406 |
| 40 | **Fusicatenibacter saccharivorans** | 432 | 391 |
| 41 | **[Clostridium] clostridioforme** | 76 | 379 |
| 42 | **Megasphaera elsdenii** | 0 | 376 |
| 43 | **Bacteroides nordii** | 56 | 369 |
| 44 | **Faecalibacterium sp. CAG:82** | 296 | 362 |
| 45 | **[Ruminococcus] gnavus** | 367 | 361 |
| 46 | **Ruminococcus sp. CAG:177** | 427 | 359 |
| 47 | **Dorea longicatena** | 402 | 358 |
| 48 | **Mitsuokella multacida** | 0 | 356 |
| 49 | **Blautia wexlerae** | 738 | 355 |
| 50 | **Eubacterium sp. CAG:251** | 0 | 351 |
| 51 | **[Ruminococcus] torques** | 790 | 342 |
| 52 | **Prevotella multisaccharivorax** | 72 | 334 |
| 53 | **Odoribacter splanchnicus** | 417 | 330 |
| 54 | **Firmicutes bacterium CAG:176** | 653 | 329 |
| 55 | **Dialister sp. CAG:486** | 143 | 328 |
| 56 | **Gemmiger formicilis** | 481 | 320 |
| 57 | **Firmicutes bacterium CAG:41** | 334 | 311 |
| 58 | **Streptococcus thermophilus** | 0 | 305 |
| 59 | **Romboutsia timonensis** | 171 | 299 |
| 60 | **Subdoligranulum sp. 60_17** | 438 | 296 |
| 61 | **Bacteroides oleiciplenus** | 0 | 286 |
| 62 | **Subdoligranulum variabile** | 275 | 281 |
| 63 | **Bacteroides plebeius** | 0 | 280 |
| 64 | **uncultured Faecalibacterium sp.** | 168 | 280 |
| 65 | **Roseburia hominis** | 94 | 273 |
| 66 | **Anaerostipes hadrus** | 350 | 266 |
| 67 | **Alistipes putredinis** | 568 | 266 |
| 68 | **butyrate-producing bacterium SS3/4** | 0 | 259 |
| 69 | **Firmicutes bacterium CAG:103** | 872 | 255 |
| 70 | **Eggerthella lenta** | 392 | 254 |
| 71 | **Catenibacterium sp. CAG:290** | 324 | 240 |
| 72 | **Mycoplasma sp. CAG:956** | 229 | 238 |
| 73 | **Veillonella dispar** | 0 | 238 |
| 74 | **Oscillibacter sp. ER4** | 283 | 238 |
| 75 | **Clostridium sp. CAG:81** | 0 | 236 |
| 76 | **Collinsella aerofaciens** | 715 | 236 |
| 77 | **Bacteroides timonensis** | 0 | 235 |
| 78 | **Turicibacter sanguinis** | 0 | 233 |
| 79 | **Bifidobacterium sp. N5G01** | 550 | 230 |
| 80 | **Bacteroides caccae** | 548 | 224 |
| 81 | **Clostridiales bacterium KLE1615** | 0 | 216 |
| 82 | **uncultured Ruminococcus sp.** | 760 | 213 |
| 83 | **Bacteroides cellulosilyticus** | 0 | 213 |
| 84 | **Clostridium sp. CAG:7** | 55 | 213 |
| 85 | **Parabacteroides merdae** | 40 | 211 |
| 86 | **Clostridium sp. CAG:389** | 291 | 208 |
| 87 | **Blautia sp. CAG:37** | 162 | 208 |
| 88 | **Coprococcus eutactus** | 158 | 200 |
| 89 | **Veillonella atypica** | 0 | 200 |
| 90 | **Ruminococcus callidus** | 32 | 200 |
| 91 | **Holdemanella biformis** | 585 | 199 |
| 92 | **Lachnospiraceae bacterium TF01-11** | 0 | 199 |
| 93 | **uncultured Lachnospira sp.** | 0 | 190 |
| 94 | **Bacteroides stercoris** | 0 | 187 |
| 95 | **Bifidobacterium catenulatum** | 142 | 183 |
| 96 | **[Eubacterium] hallii** | 346 | 174 |
| 97 | **Prevotella sp. CAG:520** | 0 | 173 |
| 98 | **Roseburia sp. CAG:18** | 283 | 173 |
| 99 | **Alistipes sp. HGB5** | 27 | 172 |
| 100 | **Haemophilus parainfluenzae** | 52 | 172 |
| 101 | **Streptococcus salivarius** | 71 | 169 |
| 102 | **Coprococcus catus** | 49 | 167 |
| 103 | **Firmicutes bacterium CAG:65** | 89 | 164 |
| 104 | **Bacteroides plebeius CAG:211** | 0 | 163 |
| 105 | **Ruminococcus bromii** | 158 | 163 |
| 106 | **Megasphaera massiliensis** | 12 | 161 |
| 107 | **Enterobacter hormaechei** | 47 | 158 |
| 108 | **Roseburia intestinalis CAG:13** | 0 | 153 |
| 109 | **Catenibacterium mitsuokai** | 198 | 149 |
| 110 | **Dakarella massiliensis** | 97 | 147 |
| 111 | **Chlamydia trachomatis** | 132 | 146 |
| 112 | **Enterobacter cloacae** | 70 | 145 |
| 113 | **Bifidobacterium pseudocatenulatum** | 251 | 145 |
| 114 | **Clostridiales bacterium VE202-06** | 0 | 144 |
| 115 | **Firmicutes bacterium CAG:65_45_313** | 0 | 142 |
| 116 | **[Clostridium] symbiosum** | 0 | 140 |
| 117 | **Veillonella parvula** | 0 | 139 |
| 118 | **Clostridiales bacterium 1_7_47FAA** | 0 | 136 |
| 119 | **Burkholderiales bacterium 1_1_47** | 0 | 135 |
| 120 | **Roseburia sp. CAG:18_43_25** | 158 | 134 |
| 121 | **Bacteroides sp. HPS0048** | 64 | 132 |
| 122 | **Weissella confusa** | 124 | 132 |
| 123 | **Clostridium sp. CAG:122** | 0 | 131 |
| 124 | **Firmicutes bacterium CAG:424** | 146 | 131 |
| 125 | **Prevotella stercorea** | 0 | 127 |
| 126 | **Ruminococcus sp. CAG:254** | 445 | 127 |
| 127 | **Bacteroides sp. 2_2_4** | 64 | 127 |
| 128 | **Enterococcus asini** | 0 | 126 |
| 129 | **Blautia sp. KLE 1732** | 185 | 126 |
| 130 | **Clostridioides difficile** | 107 | 125 |
| 131 | **Firmicutes bacterium CAG:129_59_24** | 164 | 124 |
| 132 | **[Eubacterium] siraeum** | 50 | 124 |
| 133 | **Clostridiales bacterium 42_27** | 184 | 120 |
| 134 | **Bifidobacterium kashiwanohense** | 125 | 117 |
| 135 | **Roseburia inulinivorans** | 482 | 117 |
| 136 | **Prevotella sp. CAG:732** | 28 | 114 |
| 137 | **Firmicutes bacterium CAG:170** | 235 | 113 |
| 138 | **Clostridium sp. CAG:12237_41** | 0 | 113 |
| 139 | **Clostridium bolteae CAG:59** | 0 | 113 |
| 140 | **Clostridium sp. ATCC BAA-442** | 67 | 113 |
| 141 | **Lactobacillus ruminis CAG:367** | 87 | 113 |
| 142 | **Firmicutes bacterium CAG:102** | 0 | 111 |
| 143 | **Ruminococcus sp. 5_1_39BFAA** | 166 | 105 |
| 144 | **Firmicutes bacterium CAG:114** | 480 | 103 |
| 145 | **Alistipes senegalensis** | 628 | 102 |
| 146 | **Alistipes finegoldii** | 41 | 101 |
| 147 | **Adlercreutzia equolifaciens** | 405 | 101 |
| 148 | **Sutterella sp. CAG:351** | 132 | 100 |
| 149 | **Parabacteroides distasonis** | 67 | 100 |
| 150 | **Parasutterella excrementihominis** | 0 | 98 |
| 151 | **uncultured Eubacterium sp.** | 294 | 98 |
| 152 | **Enterococcus avium** | 292 | 97 |
| 153 | **Bacteroides dorei** | 235 | 97 |
| 154 | **Bacteroides stercorirosoris** | 0 | 97 |
| 155 | **Collinsella sp. CAG:166** | 156 | 96 |
| 156 | **uncultured bacterium** | 89 | 95 |
| 157 | **Lactococcus garvieae** | 0 | 95 |
| 158 | **Prevotella sp. CAG:474** | 21 | 95 |
| 159 | **Terrisporobacter glycolicus** | 0 | 94 |
| 160 | **Oscillibacter sp. 57_20** | 66 | 94 |
| 161 | **Veillonella sp. DORA_A_3_16_22** | 0 | 94 |
| 162 | **Ruminococcus sp. CAG:90** | 118 | 92 |
| 163 | **Prevotella sp. CAG:592** | 0 | 92 |
| 164 | **Lachnospira pectinoschiza** | 0 | 92 |
| 165 | **Bacteroides intestinalis** | 0 | 89 |
| 166 | **Bacteroides caccae CAG:21** | 231 | 88 |
| 167 | **Bacteroides sp. 3_1_23** | 12 | 87 |
| 168 | **Clostridiales bacterium 41_21_two_genomes** | 21 | 87 |
| 169 | **Faecalibacterium sp. CAG:82-related_59_9** | 74 | 85 |
| 170 | **Klebsiella michiganensis** | 0 | 85 |
| 171 | **Bifidobacterium sp. N4G05** | 217 | 83 |
| 172 | **Proteobacteria bacterium CAG:139** | 0 | 81 |
| 173 | **Clostridiales bacterium 41_12_two_minus** | 83 | 81 |
| 174 | **Eubacterium sp. CAG:248** | 0 | 80 |
| 175 | **Dorea formicigenerans** | 292 | 80 |
| 176 | **Megasphaera sp. DISK 18** | 0 | 79 |
| 177 | **Coprobacillus sp. CAG:235** | 197 | 79 |
| 178 | **Prevotella stercorea CAG:629** | 0 | 79 |
| 179 | **Clostridiales bacterium 52_15** | 218 | 79 |
| 180 | **Clostridiales bacterium 59_14** | 133 | 77 |
| 181 | **Blautia sp. CAG:237** | 163 | 77 |
| 182 | **uncultured Bacteroides sp.** | 0 | 76 |
| 183 | **Alistipes finegoldii CAG:68** | 0 | 75 |
| 184 | **Dorea sp. CAG:105** | 39 | 74 |
| 185 | **Ruminococcus obeum CAG:39** | 163 | 73 |
| 186 | **Bacteroides cellulosilyticus CAG:158** | 0 | 70 |
| 187 | **Ruminococcus sp. CAG:17** | 170 | 69 |
| 188 | **Eubacterium sp. CAG:38** | 0 | 69 |
| 189 | **Clostridium sp. CAG:91** | 0 | 69 |
| 190 | **Klebsiella oxytoca** | 0 | 68 |
| 191 | **Bacteroides sp. 43_46** | 134 | 68 |
| 192 | **Prevotella sp. P3-122** | 0 | 68 |
| 193 | **Roseburia sp. CAG:471** | 101 | 68 |
| 194 | **Agathobaculum desmolans** | 54 | 67 |
| 195 | **Sutterella parvirubra** | 0 | 66 |
| 196 | **Eubacterium rectale CAG:36** | 35 | 66 |
| 197 | **Collinsella sp. 4_8_47FAA** | 144 | 65 |
| 198 | **Blautia sp. Marseille-P3201T** | 68 | 65 |
| 199 | **Bacteroides sp. 4_1_36** | 75 | 63 |
| 200 | **Odoribacter sp. 43_10** | 61 | 62 |
| 201 | **Parasutterella excrementihominis CAG:233** | 0 | 60 |
| 202 | **Bilophila sp. 4_1_30** | 0 | 60 |
| 203 | **Eubacterium eligens CAG:72** | 0 | 60 |
| 204 | **Parabacteroides merdae CAG:48** | 0 | 59 |
| 205 | **Blautia sp. CAG:37_48_57** | 87 | 59 |
| 206 | **Bifidobacterium breve** | 485 | 58 |
| 207 | **Ruminococcus sp. CAG:108** | 45 | 58 |
| 208 | **Firmicutes bacterium CAG:83** | 430 | 57 |
| 209 | **Bacteroides sp. 41_26** | 0 | 57 |
| 210 | **Anaerostipes caccae** | 0 | 56 |
| 211 | **uncultured Flavonifractor sp.** | 65 | 56 |
| 212 | **Enterococcus sp. HMSC05C03** | 31 | 56 |
| 213 | **Butyricicoccus sp. BB10** | 347 | 56 |
| 214 | **Alistipes putredinis CAG:67** | 57 | 55 |
| 215 | **Bacteroides sp. D20** | 18 | 54 |
| 216 | **Lachnoclostridium sp. An196** | 34 | 54 |
| 217 | **Dorea longicatena CAG:42** | 29 | 53 |
| 218 | **Klebsiella variicola** | 0 | 53 |
| 219 | **Akkermansia muciniphila CAG:154** | 21 | 48 |
| 220 | **Streptococcus pneumoniae** | 36 | 48 |
| 221 | **Firmicutes bacterium CAG:129** | 73 | 48 |
| 222 | **Eubacterium sp. 45_250** | 0 | 48 |
| 223 | **Bacteroides sp. D22** | 51 | 47 |
| 224 | **Eubacterium sp. CAG:76** | 0 | 47 |
| 225 | **Clostridiales bacterium VE202-28** | 0 | 47 |
| 226 | **Oscillibacter sp. CAG:241** | 69 | 46 |
| 227 | **Blautia massiliensis** | 69 | 46 |
| 228 | **Blautia sp. SF-50** | 77 | 46 |
| 229 | **Parabacteroides sp. D13** | 39 | 45 |
| 230 | **Enterococcus faecium** | 844 | 45 |
| 231 | **Mitsuokella jalaludinii** | 0 | 45 |
| 232 | **Sutterella wadsworthensis** | 0 | 44 |
| 233 | **Clostridiales bacterium 36_14** | 48 | 44 |
| 234 | **Coprococcus sp. CAG:131** | 66 | 44 |
| 235 | **Lachnoclostridium sp. An14** | 0 | 43 |
| 236 | **Lachnospiraceae bacterium 7_1_58FAA** | 18 | 43 |
| 237 | **Blautia sp. Marseille-P2398** | 86 | 43 |
| 238 | **Eubacterium hallii CAG:12** | 85 | 42 |
| 239 | **Bacteroides sp. 1_1_30** | 49 | 42 |
| 240 | **Clostridiales bacterium VE202-03** | 25 | 41 |
| 241 | **Acholeplasma sp. CAG:878** | 0 | 40 |
| 242 | **Blautia sp. CAG:52** | 175 | 40 |
| 243 | **Veillonella sp. HPA0037** | 0 | 40 |
| 244 | **Alistipes indistinctus** | 91 | 39 |
| 245 | **Ruminococcus sp. SR1/5** | 57 | 39 |
| 246 | **Roseburia sp. CAG:50** | 0 | 38 |
| 247 | **Erwinia phage vB_EamM_Y3** | 0 | 38 |
| 248 | **Bacteroides sp. D2** | 0 | 38 |
| 249 | **Eubacterium ramulus** | 34 | 37 |
| 250 | **Veillonella sp. oral taxon 158** | 0 | 37 |
| 251 | **Clostridia bacterium UC5.1-2H11** | 22 | 36 |
| 252 | **Turicibacter sp. H121** | 0 | 36 |
| 253 | **Fusobacterium sp. CAG:815** | 0 | 36 |
| 254 | **Alistipes sp. 58_9_plus** | 0 | 35 |
| 255 | **Prevotella sp. CAG:873** | 0 | 34 |
| 256 | **Clostridium sp. CAG:448** | 0 | 34 |
| 257 | **Clostridium sp. CAG:492** | 0 | 34 |
| 258 | **Firmicutes bacterium CAG:822** | 0 | 34 |
| 259 | **Veillonella sp. ACP1** | 0 | 34 |
| 260 | **Bacteroides sp. 14(A)** | 0 | 33 |
| 261 | **Clostridiales bacterium NK3B98** | 0 | 33 |
| 262 | **Ruminococcus sp. CAG:108-related_41_35** | 25 | 33 |
| 263 | **Bacteroides intestinalis CAG:315** | 0 | 32 |
| 264 | **Lactobacillus rogosae** | 0 | 32 |
| 265 | **Alistipes timonensis** | 77 | 32 |
| 266 | **Anaerotruncus sp. CAG:390** | 26 | 31 |
| 267 | **Prevotella sp. P4-65** | 0 | 31 |
| 268 | **Blautia sp. An81** | 27 | 31 |
| 269 | **Bacteroides fragilis CAG:558** | 57 | 31 |
| 270 | **Tyzzerella nexilis** | 0 | 30 |
| 271 | **Romboutsia ilealis** | 0 | 30 |
| 272 | **[Clostridium] citroniae** | 0 | 29 |
| 273 | **Prevotella sp. P5-108** | 0 | 29 |
| 274 | **Prevotella sp. P4-76** | 0 | 29 |
| 275 | **Acidiphilium sp. CAG:727** | 27 | 29 |
| 276 | **Muribaculum intestinale** | 0 | 29 |
| 277 | **Eubacterium sp. 41_20** | 27 | 29 |
| 278 | **Shigella sonnei** | 0 | 28 |
| 279 | **Terrisporobacter othiniensis** | 0 | 28 |
| 280 | **Bifidobacterium adolescentis CAG:119** | 0 | 28 |
| 281 | **Veillonella sp. ICM51a** | 0 | 28 |
| 282 | **Prevotella sp. AGR2160** | 0 | 27 |
| 283 | **Bacteroides sp. 3_1_13** | 11 | 27 |
| 284 | **Prevotella sp. CAG:279** | 320 | 27 |
| 285 | **Intestinimonas butyriciproducens** | 72 | 27 |
| 286 | **Prevotella bryantii** | 0 | 27 |
| 287 | **Streptococcus parasanguinis** | 12 | 27 |
| 288 | **Clostridium sp. CAG:43** | 0 | 27 |
| 289 | **Klebsiella aerogenes** | 503 | 27 |
| 290 | **Ruminococcus sp. CAG:330** | 37 | 26 |
| 291 | **Clostridium sp. SS2/1** | 43 | 26 |
| 292 | **uncultured Oscillibacter sp.** | 53 | 26 |
| 293 | **Prevotella sp. P5-64** | 0 | 26 |
| 294 | **Firmicutes bacterium CAG:341** | 281 | 25 |
| 295 | **Oscillibacter sp. CAG:241_62_21** | 32 | 25 |
| 296 | **Megasphaera sp. MJR8396C** | 0 | 25 |
| 297 | **Coprococcus sp. CAG:131-related_45_246** | 0 | 25 |
| 298 | **Ruminococcus sp. CAG:9** | 61 | 25 |
| 299 | **Prevotella sp. P5-60** | 0 | 25 |
| 300 | **Bacteroides sp. CAG:927** | 0 | 24 |
| 301 | **Megasphaera sp. BL7** | 0 | 24 |
| 302 | **Bacteroidales bacterium 52_46** | 0 | 24 |
| 303 | **Odoribacter splanchnicus CAG:14** | 50 | 24 |
| 304 | **Bacteroides sp. 43_108** | 0 | 24 |
| 305 | **Citrobacter koseri** | 0 | 24 |
| 306 | **Clostridium sp. CAG:221** | 0 | 23 |
| 307 | **Veillonella tobetsuensis** | 0 | 23 |
| 308 | **Collinsella sp. TF06-26** | 144 | 23 |
| 309 | **Prevotella sp. P2-180** | 0 | 23 |
| 310 | **Bacteroides intestinalis CAG:564** | 0 | 23 |
| 311 | **Lachnoclostridium edouardi** | 0 | 23 |
| 312 | **Eggerthella sp. 1_3_56FAA** | 25 | 22 |
| 313 | **Klebsiella sp. MS 92-3** | 24 | 22 |
| 314 | **Ruminococcus faecis** | 39 | 22 |
| 315 | **Bacteroides sp. 3_1_19** | 0 | 22 |
| 316 | **Bacteroides stercoris CAG:120** | 0 | 22 |
| 317 | **Bacteroides sp. 1_1_14** | 170 | 21 |
| 318 | **Parabacteroides johnsonii** | 0 | 21 |
| 319 | **Weissella cibaria** | 0 | 21 |
| 320 | **Prevotella sp. P4-67** | 0 | 21 |
| 321 | **Intestinibacter bartlettii** | 0 | 21 |
| 322 | **Lachnospiraceae bacterium 6_1_63FAA** | 21 | 21 |
| 323 | **Eubacterium siraeum CAG:80** | 0 | 21 |
| 324 | **Salmonella enterica** | 0 | 21 |
| 325 | **Prevotella sp. P4-51** | 0 | 21 |
| 326 | **Clostridium botulinum** | 0 | 20 |
| 327 | **Clostridium nexile CAG:348** | 0 | 20 |
| 328 | **Prevotellamassilia timonensis** | 0 | 20 |
| 329 | **Prevotella sp. P5-125** | 0 | 20 |
| 330 | **Clostridiales bacterium VE202-09** | 0 | 20 |
| 331 | **Shigella flexneri** | 0 | 20 |
| 332 | **Eubacterium sp. CAG76_36_125** | 0 | 20 |
| 333 | **Prevotella sp. P5-119** | 0 | 20 |
| 334 | **Prevotella ruminicola** | 0 | 19 |
| 335 | **Prevotella lascolaii** | 0 | 19 |
| 336 | **Prevotella buccae** | 0 | 19 |
| 337 | **Clostridium butyricum** | 0 | 19 |
| 338 | **Blautia sp. An46** | 20 | 19 |
| 339 | **Eggerthella sp. HGA1** | 19 | 19 |
| 340 | **Coprococcus comes** | 85 | 18 |
| 341 | **Prevotella sp. CAG:1185** | 0 | 18 |
| 342 | **Lachnospiraceae bacterium 5_1_63FAA** | 35 | 18 |
| 343 | **Bifidobacterium ruminantium** | 263 | 18 |
| 344 | **Bacteroides finegoldii** | 0 | 18 |
| 345 | **Bacteroides sp. 4_3_47FAA** | 0 | 17 |
| 346 | **Bacteroides sp. 3_1_40A** | 0 | 17 |
| 347 | **Bacteroides sp. CAG:530** | 0 | 17 |
| 348 | **Lachnospiraceae bacterium CAG:364** | 15 | 16 |
| 349 | **Prevotella sp. CAG:1124** | 0 | 16 |
| 350 | **Bacteroides vulgatus CAG:6** | 0 | 16 |
| 351 | **Prevotella baroniae** | 0 | 16 |
| 352 | **Anaerotignum lactatifermentans** | 0 | 16 |
| 353 | **Blautia hansenii** | 14 | 15 |
| 354 | **Prevotella sp. CAG:487** | 0 | 15 |
| 355 | **Prevotella timonensis** | 0 | 15 |
| 356 | **Ruminococcus gnavus CAG:126** | 17 | 15 |
| 357 | **Megasphaera sp. NM10** | 0 | 15 |
| 358 | **Prevotella buccalis** | 0 | 14 |
| 359 | **Prevotella sp. P5-92** | 0 | 14 |
| 360 | **Streptococcus infantarius** | 0 | 14 |
| 361 | **Bacteroides sp. AR20** | 0 | 14 |
| 362 | **Enterobacter sp. BIDMC 29** | 0 | 14 |
| 363 | **Lachnospiraceae bacterium 2_1_58FAA** | 13 | 14 |
| 364 | **Bacteroides uniformis CAG:3** | 18 | 14 |
| 365 | **Prevotella sp. CAG:5226** | 0 | 14 |
| 366 | **Bacteroides sartorii** | 0 | 14 |
| 367 | **Blautia schinkii** | 0 | 14 |
| 368 | **[Clostridium] lavalense** | 0 | 13 |
| 369 | **Prevotella intermedia** | 0 | 13 |
| 370 | **Bacteroides mediterraneensis** | 0 | 13 |
| 371 | **Veillonella sp. 6_1_27** | 0 | 13 |
| 372 | **Lactococcus lactis** | 140 | 13 |
| 373 | **Bacteroides faecis** | 0 | 12 |
| 374 | **Lachnospiraceae bacterium JC7** | 37 | 12 |
| 375 | **Prevotella histicola** | 0 | 11 |
| 376 | **Sutterella sp. KLE1602** | 0 | 11 |
| 377 | **Prevotella oralis** | 0 | 11 |
| 378 | **Bifidobacterium bifidum CAG:234** | 0 | 10 |
| 379 | **Prevotella sp. P4-119** | 0 | 10 |
| 380 | **Prevotella paludivivens** | 0 | 10 |
| 381 | **Prevotella sp. tc2-28** | 0 | 10 |
| 382 | **Prevotella sp. P5-126** | 0 | 10 |
| 383 | **Prevotella sp. 109** | 0 | 10 |
| 384 | **Prevotella brevis** | 0 | 10 |
| 385 | **Prevotella oris** | 0 | 9 |
| 386 | **Prevotella sp. DNF00663** | 0 | 9 |
| 387 | **Prevotella oryzae** | 0 | 9 |
| 388 | **Prevotella sp. CAG:255** | 0 | 9 |
| 389 | **Prevotella sp. S7-1-8** | 0 | 9 |
| 390 | **Prevotella dentalis** | 0 | 9 |
| 391 | **Prevotella sp. KH2C16** | 0 | 9 |
| 392 | **Prevotella sp. CAG:1058** | 0 | 9 |
| 393 | **Prevotella maculosa** | 0 | 9 |
| 394 | **Prevotella bergensis** | 0 | 9 |
| 395 | **Ruminococcaceae bacterium D16** | 14 | 8 |
| 396 | **Dorea formicigenerans CAG:28** | 31 | 0 |
| 397 | **Rummeliibacillus stabekisii** | 41 | 0 |
| 398 | **Bifidobacterium pseudolongum** | 30 | 0 |
| 399 | **Clostridium sp. CAG:138** | 415 | 0 |
| 400 | **Blautia sp. CAG:257** | 39 | 0 |
| 401 | **Enterococcus sp. HMSC072H05** | 14 | 0 |
| 402 | **Bacteroides dorei CAG:222** | 22 | 0 |
| 403 | **Bacteroides sp. CAG:189** | 67 | 0 |
| 404 | **[Desulfotomaculum] guttoideum** | 17 | 0 |
| 405 | **Tissierellia bacterium S5-A11** | 21 | 0 |
| 406 | **Ruminococcus sp. CAG:382** | 21 | 0 |
| 407 | **Peptococcus niger** | 27 | 0 |
| 408 | **Oribacterium sp. C9** | 24 | 0 |
| 409 | **Olsenella provencensis** | 22 | 0 |
| 410 | **Collinsella sp. CAG:289** | 85 | 0 |
| 411 | **Ruthenibacterium lactatiformans** | 63 | 0 |
| 412 | **Acinetobacter sp. NIPH 899** | 71 | 0 |
| 413 | **Oribacterium sp. WCC10** | 33 | 0 |
| 414 | **Clostridium sp. CAG:609** | 259 | 0 |
| 415 | **Clostridium sp. CAG:571** | 33 | 0 |
| 416 | **Butyricimonas virosa** | 146 | 0 |
| 417 | **Slackia piriformis** | 52 | 0 |
| 418 | **Firmicutes bacterium CAG:110** | 278 | 0 |
| 419 | **Achromobacter xylosoxidans** | 12 | 0 |
| 420 | **Lactobacillus mucosae** | 321 | 0 |
| 421 | **Clostridium sp. CAG:433** | 54 | 0 |
| 422 | **Clostridium sp. CAG:226** | 155 | 0 |
| 423 | **Solobacterium moorei** | 27 | 0 |
| 424 | **bacterium LF-3** | 22 | 0 |
| 425 | **Anaerococcus prevotii** | 17 | 0 |
| 426 | **Olsenella sp. An188** | 24 | 0 |
| 427 | **Clostridium minihomine** | 16 | 0 |
| 428 | **Acinetobacter sp. NIPH 2171** | 133 | 0 |
| 429 | **Anaeromassilibacillus senegalensis** | 19 | 0 |
| 430 | **Dialister invisus** | 60 | 0 |
| 431 | **Firmicutes bacterium CAG:646** | 12 | 0 |
| 432 | **bacterium MS4** | 32 | 0 |
| 433 | **Gordonibacter pamelaeae** | 82 | 0 |
| 434 | **Parabacteroides sp. HGS0025** | 18 | 0 |
| 435 | **Anaeromassilibacillus sp. Marseille-P3371** | 12 | 0 |
| 436 | **Peptoniphilus senegalensis** | 37 | 0 |
| 437 | **Clostridium sp. 7_2_43FAA** | 17 | 0 |
| 438 | **Peptoniphilus duerdenii** | 16 | 0 |
| 439 | **Comamonas testosteroni** | 90 | 0 |
| 440 | **Anaeromassilibacillus sp. An200** | 9 | 0 |
| 441 | **Lysinibacillus sp. ZYM-1** | 44 | 0 |
| 442 | **Mycoplasma sp. CAG:472** | 53 | 0 |
| 443 | **Clostridium sp. L2-50** | 37 | 0 |
| 444 | **Collinsella sp. MS5** | 44 | 0 |
| 445 | **Firmicutes bacterium CAG:555** | 26 | 0 |
| 446 | **Bacillus kochii** | 12 | 0 |
| 447 | **Faecalibacterium sp. CAG:74_58_120** | 295 | 0 |
| 448 | **Clostridium sp. CAG:793** | 270 | 0 |
| 449 | **Neglecta timonensis** | 26 | 0 |
| 450 | **[Clostridium] celerecrescens** | 750 | 0 |
| 451 | **Clostridium sp. ASBs410** | 20 | 0 |
| 452 | **Enterococcus casseliflavus** | 251 | 0 |
| 453 | **Eggerthella timonensis** | 23 | 0 |
| 454 | **Faecalibacterium sp. CAG:74** | 439 | 0 |
| 455 | **Clostridiales bacterium Marseille-P2846** | 187 | 0 |
| 456 | **Anaerococcus vaginalis** | 30 | 0 |
| 457 | **Anaeromassilibacillus sp. An250** | 39 | 0 |
| 458 | **Acinetobacter sp. CIP 101934** | 13 | 0 |
| 459 | **Firmicutes bacterium CAG:24** | 145 | 0 |
| 460 | **Firmicutes bacterium HGW-Firmicutes-16** | 22 | 0 |
| 461 | **Barnesiella intestinihominis** | 121 | 0 |
| 462 | **Enterococcus gallinarum** | 214 | 0 |
| 463 | **Lysinibacillus sp. FJAT-14222** | 245 | 0 |
| 464 | **Alistipes shahii** | 40 | 0 |
| 465 | **Peptoniphilus timonensis** | 41 | 0 |
| 466 | **Akkermansia sp. CAG:344** | 85 | 0 |
| 467 | **Lagierella massiliensis** | 39 | 0 |
| 468 | **Acinetobacter sp. LCT-H3** | 26 | 0 |
| 469 | **Drancourtella massiliensis** | 12 | 0 |
| 470 | **Subdoligranulum sp. 4_3_54A2FAA** | 88 | 0 |
| 471 | **Peptoniphilus harei** | 30 | 0 |
| 472 | **Ruminococcus sp. CAG:9-related_41_34** | 11 | 0 |
| 473 | **Ruminococcus lactaris** | 22 | 0 |
| 474 | **Firmicutes bacterium CAG:24053_14** | 48 | 0 |
| 475 | **Enterorhabdus caecimuris** | 28 | 0 |
| 476 | **Alistipes sp. Marseille-P2431** | 21 | 0 |
| 477 | **Bacteroides thetaiotaomicron CAG:40** | 42 | 0 |
| 478 | **Ruminococcus flavefaciens** | 30 | 0 |
| 479 | **Blautia sp. Marseille-P3087** | 46 | 0 |
| 480 | **Oribacterium sp. P6A1** | 25 | 0 |
| 481 | **Clostridium sp. C105KSO15** | 124 | 0 |
| 482 | **Achromobacter sp. Root170** | 21 | 0 |
| 483 | **Clostridium sp. CAG:264** | 101 | 0 |
| 484 | **Lysinibacillus sphaericus** | 108 | 0 |
| 485 | **Clostridium sp. CAG:1024** | 39 | 0 |
| 486 | **Urinacoccus sp. Marseille-P3926** | 136 | 0 |
| 487 | **Enterococcus sp. FDAARGOS_375** | 30 | 0 |
| 488 | **Clostridiales bacterium** | 81 | 0 |
| 489 | **Lysinibacillus boronitolerans** | 27 | 0 |
| 490 | **Collinsella bouchesdurhonensis** | 32 | 0 |
| 491 | **Rummeliibacillus pycnus** | 562 | 0 |
| 492 | **Erysipelotrichaceae bacterium NK3D112** | 33 | 0 |
| 493 | **Collinsella sp. 60_9** | 26 | 0 |
| 494 | **Alistipes obesi** | 352 | 0 |
| 495 | **Dialister invisus CAG:218** | 290 | 0 |
| 496 | **Eubacterium sp. CAG:161** | 21 | 0 |
| 497 | **Bacteroides sp. 3_1_33FAA** | 22 | 0 |
| 498 | **Firmicutes bacterium CAG:110_56_8** | 45 | 0 |
| 499 | **uncultured Coprococcus sp.** | 83 | 0 |
| 500 | **Paraclostridium bifermentans** | 17 | 0 |
| 501 | **Monoglobus pectinilyticus** | 24 | 0 |
| 502 | **Oribacterium sp. NK2B42** | 20 | 0 |
| 503 | **Lactobacillus brevis** | 32 | 0 |
| 504 | **Senegalimassilia anaerobia** | 122 | 0 |
| 505 | **Acinetobacter baumannii** | 228 | 0 |
| 506 | **Clostridium sp. CAG:567** | 236 | 0 |
| 507 | **Coprococcus sp. ART55/1** | 22 | 0 |
| 508 | **Peptoniphilus sp. HMSC075B08** | 105 | 0 |
| 509 | **Enterococcus pallens** | 17 | 0 |
| 510 | **Finegoldia magna** | 82 | 0 |
| 511 | **Lysinibacillus sp. FJAT-14745** | 97 | 0 |
| 512 | **Coprobacillus sp. 8_1_38FAA** | 10 | 0 |
| 513 | **Peptoniphilus sp. oral taxon 375** | 118 | 0 |
| 514 | **uncultured crAssphage** | 31 | 0 |
| 515 | **Gordonibacter massiliensis** | 20 | 0 |
| 516 | **Olsenella sp. An290** | 19 | 0 |
| 517 | **Peptoniphilus grossensis** | 71 | 0 |
| 518 | **Bacteroides sp. 9_1_42FAA** | 31 | 0 |
| 519 | **Firmicutes bacterium CAG:176_63_11** | 30 | 0 |
| 520 | **Enterococcus faecalis** | 238 | 0 |
| 521 | **Lactobacillus plantarum** | 259 | 0 |
| 522 | **Lysinibacillus fusiformis** | 42 | 0 |
| 523 | **Eubacterium sp. 38_16** | 21 | 0 |
| 524 | **Peptoniphilus sp. HMSC062D09** | 42 | 0 |
| 525 | **Alistipes sp. CAG:53** | 39 | 0 |
| 526 | **Bacteroidales bacterium 43_8** | 22 | 0 |
| 527 | **Peptoniphilus phoceensis** | 55 | 0 |
| 528 | **Peptoniphilus sp. BV3AC2** | 12 | 0 |
| 529 | **Clostridium sp. CAG:302** | 267 | 0 |
| 530 | **Gordonibacter urolithinfaciens** | 567 | 0 |
| 531 | **Acinetobacter sp. YZS-X1-1** | 115 | 0 |
| 532 | **Lysinibacillus xylanilyticus** | 242 | 0 |
| 533 | **Acinetobacter schindleri** | 91 | 0 |
| 534 | **Bacteroides sp. CAG:20** | 60 | 0 |
| 535 | **Clostridium sp. CAG:269** | 67 | 0 |
| 536 | **Alistipes sp. cv1** | 20 | 0 |
| 537 | **Roseburia inulinivorans CAG:15** | 111 | 0 |
| 538 | **Urmitella timonensis** | 24 | 0 |
| 539 | **Olsenella sp. An293** | 20 | 0 |
| 540 | **Eisenbergiella tayi** | 73 | 0 |
| 541 | **Enterococcus saccharolyticus** | 16 | 0 |
| 542 | **Parabacteroides goldsteinii** | 602 | 0 |
| 543 | **Marvinbryantia formatexigens** | 22 | 0 |
| 544 | **Lysinibacillus macroides** | 49 | 0 |
| 545 | **Bacteroides salyersiae** | 248 | 0 |
| 546 | **Eubacterium sp. CAG:146** | 52 | 0 |
| 547 | **Peptoniphilus coxii** | 318 | 0 |
| 548 | **Libanicoccus massiliensis** | 25 | 0 |
| 549 | **Roseburia sp. CAG:182** | 36 | 0 |
| 550 | **Clostridium sp. CAG:413** | 26 | 0 |
| 551 | **Dorea sp. AGR2135** | 24 | 0 |
| 552 | **Acinetobacter lwoffii** | 12 | 0 |

**Table 4: Species level: Mean abundance at baseline and post-intervention in**

**Gr.2 (Nichi Glucan)**

| **S.No** | **Species** | **Mean abundance** | |
| --- | --- | --- | --- |
|  |  | **Baseline** | **Post-Intervention** |
| 1 | **Faecalibacterium prausnitzii** | 3613 | 6596 |
| 2 | **Bifidobacterium longum** | 2012 | 1446 |
| 3 | **Firmicutes bacterium CAG:124** | 1684 | 1308 |
| 4 | **Trichosporon asahii** | 1683 | 0 |
| 5 | **Bifidobacterium adolescentis** | 1376 | 931 |
| 6 | **Escherichia coli** | 1264 | 471 |
| 7 | **uncultured Clostridium sp.** | 1062 | 2726 |
| 8 | **Collinsella aerofaciens** | 1048 | 729 |
| 9 | **uncultured Blautia sp.** | 981 | 879 |
| 10 | **Ruminococcus sp. CAG:177** | 977 | 565 |
| 11 | **Firmicutes bacterium CAG:103** | 972 | 925 |
| 12 | **Bacteroides fragilis** | 960 | 1186 |
| 13 | **Blautia obeum** | 877 | 777 |
| 14 | **Gemmiger formicilis** | 865 | 1022 |
| 15 | **Firmicutes bacterium CAG:170** | 847 | 989 |
| 16 | **Dorea longicatena** | 819 | 785 |
| 17 | **Firmicutes bacterium CAG:110** | 787 | 604 |
| 18 | **Clostridium sp. CAG:226** | 779 | 321 |
| 19 | **Clostridium sp. CAG:138** | 743 | 528 |
| 20 | **uncultured Ruminococcus sp.** | 726 | 913 |
| 21 | **Bacteroides uniformis** | 707 | 512 |
| 22 | **Alistipes sp. CAG:435** | 690 | 721 |
| 23 | **Hungatella hathewayi** | 681 | 171 |
| 24 | **Holdemanella biformis** | 649 | 546 |
| 25 | **Oscillibacter sp. CAG:241** | 634 | 419 |
| 26 | **Firmicutes bacterium CAG:176** | 617 | 945 |
| 27 | **Firmicutes bacterium CAG:83** | 612 | 400 |
| 28 | **Subdoligranulum sp. 60_17** | 588 | 734 |
| 29 | **Pichia kudriavzevii** | 582 | 0 |
| 30 | **Bacteroides thetaiotaomicron** | 581 | 562 |
| 31 | **Prevotella copri** | 568 | 1458 |
| 32 | **Eubacterium sp. CAG:202** | 536 | 0 |
| 33 | **Enterococcus faecium** | 528 | 0 |
| 34 | **Klebsiella pneumoniae** | 524 | 44 |
| 35 | **Rummeliibacillus pycnus** | 516 | 0 |
| 36 | **Senegalimassilia anaerobia** | 505 | 56 |
| 37 | **[Eubacterium] rectale** | 499 | 1165 |
| 38 | **Firmicutes bacterium CAG:114** | 489 | 386 |
| 39 | **Bifidobacterium bifidum** | 476 | 418 |
| 40 | **Bacteroides ovatus** | 472 | 448 |
| 41 | **Bacteroides vulgatus** | 467 | 375 |
| 42 | **Blautia wexlerae** | 464 | 486 |
| 43 | **Lactobacillus ruminis** | 464 | 634 |
| 44 | **[Ruminococcus] torques** | 452 | 347 |
| 45 | **Fusicatenibacter saccharivorans** | 450 | 624 |
| 46 | **[Eubacterium] hallii** | 442 | 218 |
| 47 | **Olsenella umbonata** | 441 | 37 |
| 48 | **Desulfovibrio piger** | 434 | 303 |
| 49 | **Dialister sp. CAG:486** | 433 | 447 |
| 50 | **uncultured Eubacterium sp.** | 429 | 294 |
| 51 | **Enterococcus avium** | 425 | 0 |
| 52 | **Faecalibacterium sp. CAG:74** | 415 | 485 |
| 53 | **Bacteroides caccae** | 398 | 152 |
| 54 | **Oscillibacter sp. CAG:241_62_21** | 392 | 469 |
| 55 | **Romboutsia timonensis** | 388 | 112 |
| 56 | **Eubacterium sp. CAG:180** | 379 | 338 |
| 57 | **Parabacteroides merdae** | 377 | 101 |
| 58 | **Pediococcus pentosaceus** | 371 | 0 |
| 59 | **Enterococcus faecalis** | 362 | 0 |
| 60 | **Clostridiales bacterium Marseille-P2846** | 358 | 355 |
| 61 | **Clostridium sp. CAG:221** | 355 | 326 |
| 62 | **Butyricicoccus sp. BB10** | 334 | 177 |
| 63 | **[Clostridium] celerecrescens** | 333 | 0 |
| 64 | **Lentisphaerae bacterium GWF2_44_16** | 332 | 307 |
| 65 | **[Clostridium] bolteae** | 331 | 43 |
| 66 | **Clostridiales bacterium 42_27** | 329 | 463 |
| 67 | **Prevotella sp. CAG:279** | 326 | 754 |
| 68 | **Bifidobacterium sp. N5G01** | 325 | 383 |
| 69 | **Firmicutes bacterium CAG:129** | 324 | 380 |
| 70 | **Cloacibacillus porcorum** | 322 | 248 |
| 71 | **Clostridium sp. CAG:1024** | 319 | 488 |
| 72 | **Prevotella copri CAG:164** | 318 | 771 |
| 73 | **Catenibacterium mitsuokai** | 312 | 274 |
| 74 | **Eggerthella lenta** | 309 | 60 |
| 75 | **Mycoplasma sp. CAG:956** | 306 | 117 |
| 76 | **Oscillibacter sp. ER4** | 302 | 587 |
| 77 | **Roseburia faecis** | 301 | 662 |
| 78 | **Coraliomargarita sp. CAG:312** | 292 | 338 |
| 79 | **Clostridiales bacterium 52_15** | 290 | 332 |
| 80 | **Catenibacterium sp. CAG:290** | 287 | 180 |
| 81 | **Pediococcus acidilactici** | 282 | 0 |
| 82 | **Bacteroides dorei** | 281 | 357 |
| 83 | **Clostridiales bacterium 59_14** | 280 | 386 |
| 84 | **Firmicutes bacterium CAG:176_63_11** | 280 | 400 |
| 85 | **Alistipes putredinis** | 278 | 201 |
| 86 | **Odoribacter splanchnicus** | 277 | 129 |
| 87 | **bacterium OL-1** | 275 | 15 |
| 88 | **Parabacteroides sp. SN4** | 274 | 249 |
| 89 | **[Eubacterium] eligens** | 273 | 552 |
| 90 | **Alistipes indistinctus** | 272 | 13 |
| 91 | **Alistipes obesi** | 272 | 0 |
| 92 | **Clostridium sp. CAG:510** | 270 | 449 |
| 93 | **Dialister sp. CAG:357** | 270 | 360 |
| 94 | **Faecalibacterium sp. CAG:74_58_120** | 264 | 368 |
| 95 | **Lactobacillus brevis** | 262 | 0 |
| 96 | **Bifidobacterium ruminantium** | 261 | 257 |
| 97 | **Anaerostipes hadrus** | 260 | 422 |
| 98 | **Enterococcus casseliflavus** | 259 | 0 |
| 99 | **Cloacibacillus sp. An23** | 257 | 0 |
| 100 | **Bacteroides plebeius** | 250 | 250 |
| 101 | **Weissella confusa** | 249 | 12 |
| 102 | **Lactobacillus plantarum** | 249 | 0 |
| 103 | **Bacteroides sp. CAG:545** | 246 | 379 |
| 104 | **Clostridium sp. CAG:452** | 245 | 120 |
| 105 | **uncultured Faecalibacterium sp.** | 243 | 437 |
| 106 | **Collinsella sp. 4_8_47FAA** | 239 | 166 |
| 107 | **Firmicutes bacterium CAG:555** | 237 | 281 |
| 108 | **Ruminococcus sp. CAG:724** | 236 | 367 |
| 109 | **Roseburia inulinivorans** | 233 | 1197 |
| 110 | **Lentisphaerae bacterium GWF2_45_14** | 232 | 228 |
| 111 | **Bacteroides xylanisolvens** | 232 | 285 |
| 112 | **uncultured Butyricicoccus sp.** | 230 | 995 |
| 113 | **Butyricimonas virosa** | 230 | 0 |
| 114 | **Bacteroides intestinalis** | 230 | 135 |
| 115 | **Collinsella sp. CAG:166** | 228 | 187 |
| 116 | **Ruminococcus sp. CAG:488** | 225 | 211 |
| 117 | **Coprococcus catus** | 225 | 412 |
| 118 | **Klebsiella aerogenes** | 224 | 0 |
| 119 | **Phascolarctobacterium sp. CAG:207** | 220 | 21 |
| 120 | **Acidiphilium sp. CAG:727** | 219 | 555 |
| 121 | **Parabacteroides gordonii** | 217 | 0 |
| 122 | **[Eubacterium] siraeum** | 212 | 189 |
| 123 | **Adlercreutzia equolifaciens** | 210 | 334 |
| 124 | **Butyrivibrio sp. CAG:318** | 208 | 0 |
| 125 | **Blautia sp. CAG:37** | 207 | 203 |
| 126 | **Clostridium sp. CAG:448** | 203 | 192 |
| 127 | **Bacteroides salyersiae** | 203 | 193 |
| 128 | **Coprobacillus sp. CAG:235** | 197 | 145 |
| 129 | **Clostridiales bacterium** | 194 | 147 |
| 130 | **Bifidobacterium pseudocatenulatum** | 194 | 174 |
| 131 | **Collinsella sp. TF06-26** | 189 | 155 |
| 132 | **Alistipes sp. CAG:53** | 189 | 180 |
| 133 | **Clostridium sp. CAG:571** | 189 | 14 |
| 134 | **Sutterella sp. CAG:397** | 188 | 193 |
| 135 | **Libanicoccus massiliensis** | 188 | 119 |
| 136 | **Succinatimonas sp. CAG:777** | 187 | 376 |
| 137 | **Subdoligranulum variabile** | 187 | 244 |
| 138 | **Anaerotruncus sp. CAG:390** | 185 | 275 |
| 139 | **Bifidobacterium angulatum** | 185 | 332 |
| 140 | **Angelakisella massiliensis** | 184 | 79 |
| 141 | **Alistipes senegalensis** | 183 | 57 |
| 142 | **Megamonas funiformis** | 182 | 175 |
| 143 | **Ruminococcus obeum CAG:39** | 180 | 137 |
| 144 | **Acidaminococcus fermentans** | 180 | 199 |
| 145 | **Lactobacillus mucosae** | 178 | 0 |
| 146 | **Firmicutes bacterium CAG:129_59_24** | 174 | 400 |
| 147 | **Chlamydia trachomatis** | 174 | 127 |
| 148 | **Lentisphaerae bacterium GWF2_52_8** | 172 | 173 |
| 149 | **Ruminococcus bromii** | 172 | 301 |
| 150 | **Eubacterium sp. CAG:581** | 171 | 148 |
| 151 | **Clostridium sp. CAG:433** | 170 | 8 |
| 152 | **Weissella cibaria** | 170 | 0 |
| 153 | **Methanobrevibacter smithii** | 161 | 159 |
| 154 | **Enterococcus gallinarum** | 161 | 0 |
| 155 | **Firmicutes bacterium CAG:240** | 159 | 135 |
| 156 | **Clostridium sp. CAG:302** | 159 | 20 |
| 157 | **Bacteroides plebeius CAG:211** | 158 | 160 |
| 158 | **Firmicutes bacterium CAG:24053_14** | 158 | 158 |
| 159 | **Eubacterium limosum** | 157 | 0 |
| 160 | **Clostridium sp. CAG:349** | 157 | 143 |
| 161 | **Clostridium sp. CAG:43** | 156 | 162 |
| 162 | **Parabacteroides sp. HGS0025** | 155 | 0 |
| 163 | **Akkermansia muciniphila** | 153 | 218 |
| 164 | **Clostridium sp. CAG:245** | 152 | 15 |
| 165 | **Enterococcus hirae** | 151 | 0 |
| 166 | **Duodenibacillus massiliensis** | 150 | 181 |
| 167 | **Firmicutes bacterium CAG:272** | 149 | 254 |
| 168 | **Coprococcus eutactus** | 148 | 100 |
| 169 | **Verrucomicrobia bacterium CAG:312_58_20** | 148 | 273 |
| 170 | **Faecalibacterium sp. CAG:82** | 147 | 398 |
| 171 | **Lysinibacillus xylanilyticus** | 147 | 0 |
| 172 | **Firmicutes bacterium CAG:460** | 144 | 166 |
| 173 | **Peptoniphilus coxii** | 141 | 0 |
| 174 | **uncultured bacterium** | 141 | 176 |
| 175 | **Lysinibacillus sp. FJAT-14222** | 141 | 0 |
| 176 | **Bifidobacterium breve** | 139 | 132 |
| 177 | **Clostridium sp. CAG:245_30_32** | 138 | 9 |
| 178 | **Clostridium sp. CAG:451** | 136 | 30 |
| 179 | **Sutterella wadsworthensis** | 136 | 279 |
| 180 | **Parabacteroides goldsteinii** | 135 | 13 |
| 181 | **Streptococcus mutans** | 133 | 0 |
| 182 | **Roseburia hominis** | 132 | 630 |
| 183 | **Streptococcus salivarius** | 131 | 110 |
| 184 | **Alistipes sp. CAG:514** | 131 | 147 |
| 185 | **Firmicutes bacterium CAG:137** | 131 | 72 |
| 186 | **Roseburia sp. CAG:18** | 130 | 229 |
| 187 | **Ruminococcus sp. 5_1_39BFAA** | 129 | 152 |
| 188 | **Clostridioides difficile** | 127 | 64 |
| 189 | **Flavonifractor plautii** | 125 | 221 |
| 190 | **Gordonibacter urolithinfaciens** | 125 | 0 |
| 191 | **Firmicutes bacterium CAG:110_56_8** | 123 | 87 |
| 192 | **Pyramidobacter sp. C12-8** | 122 | 0 |
| 193 | **Ruminococcus sp. CAG:17** | 122 | 151 |
| 194 | **Bifidobacterium sp. N4G05** | 119 | 125 |
| 195 | **Streptococcus thermophilus** | 118 | 0 |
| 196 | **Clostridium sp. CAG:568** | 117 | 204 |
| 197 | **Blautia sp. KLE 1732** | 115 | 51 |
| 198 | **Parabacteroides merdae CAG:48** | 114 | 30 |
| 199 | **Dorea formicigenerans** | 114 | 119 |
| 200 | **Barnesiella intestinihominis** | 112 | 77 |
| 201 | **[Clostridium] clostridioforme** | 111 | 124 |
| 202 | **Intestinimonas butyriciproducens** | 110 | 124 |
| 203 | **Olsenella scatoligenes** | 107 | 13 |
| 204 | **Clostridium sp. CAG:413** | 107 | 472 |
| 205 | **uncultured Flavonifractor sp.** | 105 | 138 |
| 206 | **Clostridium sp. CAG:343** | 104 | 0 |
| 207 | **Alistipes shahii** | 104 | 102 |
| 208 | **Pyramidobacter piscolens** | 103 | 0 |
| 209 | **Firmicutes bacterium CAG:238** | 103 | 75 |
| 210 | **Collinsella sp. CAG:289** | 102 | 0 |
| 211 | **Bacteroides caccae CAG:21** | 102 | 107 |
| 212 | **Acinetobacter baumannii** | 101 | 0 |
| 213 | **Lentisphaerae bacterium GWF2_50_93** | 100 | 109 |
| 214 | **Bacteroides sp. 2_2_4** | 97 | 83 |
| 215 | **Eubacterium sp. CAG:841** | 93 | 105 |
| 216 | **Ruminococcus sp. CAG:382** | 93 | 170 |
| 217 | **Firmicutes bacterium CAG:270** | 92 | 45 |
| 218 | **Olsenella sp. kh2p3** | 90 | 13 |
| 219 | **Phascolarctobacterium succinatutens** | 90 | 420 |
| 220 | **Lactobacillus fermentum** | 90 | 0 |
| 221 | **Mycoplasma sp. CAG:877** | 89 | 41 |
| 222 | **Bacteroides intestinalis CAG:564** | 87 | 60 |
| 223 | **Oxalobacter formigenes** | 87 | 0 |
| 224 | **Eisenbergiella tayi** | 86 | 99 |
| 225 | **Parabacteroides distasonis** | 86 | 157 |
| 226 | **Lentisphaerae bacterium GWF2_49_21** | 84 | 96 |
| 227 | **Dorea longicatena CAG:42** | 84 | 83 |
| 228 | **Ruminococcus sp. CAG:563** | 83 | 191 |
| 229 | **Olsenella sp. KH3B4** | 83 | 16 |
| 230 | **Lactobacillus ruminis CAG:367** | 82 | 106 |
| 231 | **Blautia sp. CAG:37_48_57** | 82 | 140 |
| 232 | **Alistipes sp. 56_sp_Nov_56_25** | 82 | 62 |
| 233 | **Enterococcus thailandicus** | 80 | 0 |
| 234 | **Alistipes sp. HGB5** | 80 | 0 |
| 235 | **Blautia sp. CAG:237** | 78 | 207 |
| 236 | **Bacteroides stercoris** | 77 | 19 |
| 237 | **Firmicutes bacterium HGW-Firmicutes-9** | 75 | 94 |
| 238 | **Clostridium sp. CAG:1193** | 75 | 125 |
| 239 | **Roseburia sp. CAG:18_43_25** | 75 | 173 |
| 240 | **Firmicutes bacterium HGW-Firmicutes-16** | 74 | 73 |
| 241 | **Oscillibacter sp. 57_20** | 74 | 403 |
| 242 | **Olsenella provencensis** | 74 | 7 |
| 243 | **Olsenella sp. An188** | 74 | 0 |
| 244 | **Anaerotruncus colihominis** | 73 | 71 |
| 245 | **Clostridium sp. CAG:524** | 73 | 71 |
| 246 | **Bacteroides coprocola CAG:162** | 72 | 79 |
| 247 | **Firmicutes bacterium CAG:41** | 72 | 310 |
| 248 | **Coprococcus comes** | 71 | 125 |
| 249 | **Ruminococcus sp. CAG:90** | 71 | 9 |
| 250 | **Clostridium sp. CAG:492** | 70 | 0 |
| 251 | **Pygmaiobacter massiliensis** | 69 | 0 |
| 252 | **Lysinibacillus sphaericus** | 69 | 0 |
| 253 | **Roseburia intestinalis** | 69 | 1218 |
| 254 | **Ruminococcus flavefaciens** | 69 | 267 |
| 255 | **uncultured Coprococcus sp.** | 68 | 22 |
| 256 | **Lactococcus lactis** | 68 | 0 |
| 257 | **uncultured Oscillibacter sp.** | 66 | 87 |
| 258 | **Gordonibacter pamelaeae** | 65 | 0 |
| 259 | **Bacteroides eggerthii** | 64 | 0 |
| 260 | **Bifidobacterium kashiwanohense** | 64 | 82 |
| 261 | **Clostridiales bacterium 41_12_two_minus** | 63 | 195 |
| 262 | **Collinsella sp. 60_9** | 63 | 17 |
| 263 | **Allisonella histaminiformans** | 62 | 54 |
| 264 | **Megasphaera elsdenii** | 61 | 262 |
| 265 | **Lysinibacillus sp. FJAT-14745** | 61 | 0 |
| 266 | **Ruminococcus sp. CAG:9** | 60 | 59 |
| 267 | **Urinacoccus sp. Marseille-P3926** | 60 | 0 |
| 268 | **Bacteroides coprocola** | 60 | 99 |
| 269 | **Olsenella sp. An285** | 59 | 0 |
| 270 | **Acinetobacter sp. NIPH 2171** | 59 | 0 |
| 271 | **Clostridium sp. CAG:7** | 59 | 396 |
| 272 | **Bifidobacterium catenulatum** | 58 | 75 |
| 273 | **Olsenella sp. An290** | 58 | 0 |
| 274 | **Eubacterium hallii CAG:12** | 57 | 41 |
| 275 | **Streptococcus pneumoniae** | 57 | 8 |
| 276 | **Collinsella sp. MS5** | 56 | 0 |
| 277 | **Eggerthella sp. CAG:209** | 56 | 0 |
| 278 | **Bacteroides sp. CAG:20** | 56 | 37 |
| 279 | **Clostridium sp. C105KSO15** | 55 | 0 |
| 280 | **Bacteroides sp. CAG:189** | 55 | 36 |
| 281 | **Clostridium sp. CAG:594** | 55 | 0 |
| 282 | **Bacteroides sp. AR29** | 55 | 0 |
| 283 | **Subdoligranulum sp. 4_3_54A2FAA** | 54 | 65 |
| 284 | **Ruminococcus bicirculans** | 54 | 172 |
| 285 | **Oscillibacter sp. 1-3** | 53 | 70 |
| 286 | **Blautia sp. Marseille-P3087** | 53 | 39 |
| 287 | **Turicibacter sanguinis** | 52 | 26 |
| 288 | **Peptoniphilus sp. oral taxon 375** | 52 | 0 |
| 289 | **Olsenella sp. An293** | 52 | 0 |
| 290 | **Bacteroides intestinalis CAG:315** | 51 | 32 |
| 291 | **Acinetobacter sp. YZS-X1-1** | 51 | 0 |
| 292 | **Ruthenibacterium lactatiformans** | 51 | 69 |
| 293 | **Coprococcus eutactus CAG:665** | 51 | 0 |
| 294 | **Firmicutes bacterium CAG:321** | 51 | 69 |
| 295 | **Bacteroides sp. D20** | 50 | 32 |
| 296 | **Clostridium sp. CAG:264** | 50 | 38 |
| 297 | **Blautia producta** | 49 | 0 |
| 298 | **Firmicutes bacterium CAG:176_59_8** | 49 | 90 |
| 299 | **Bacteroides sp. 1_1_14** | 48 | 18 |
| 300 | **Erysipelotrichaceae bacterium 6_1_45** | 48 | 0 |
| 301 | **Peptoniphilus sp. HMSC075B08** | 47 | 0 |
| 302 | **Selenomonas bovis** | 47 | 168 |
| 303 | **Bacteroides sp. 4_1_36** | 45 | 27 |
| 304 | **Slackia piriformis** | 44 | 0 |
| 305 | **Enterobacter cloacae** | 44 | 0 |
| 306 | **[Clostridium] citroniae** | 43 | 0 |
| 307 | **Ruminococcus sp. CAG:254** | 43 | 486 |
| 308 | **Alistipes finegoldii** | 43 | 143 |
| 309 | **Haemophilus parainfluenzae** | 43 | 87 |
| 310 | **Alistipes onderdonkii** | 42 | 14 |
| 311 | **Actinomyces sp. HPA0247** | 42 | 0 |
| 312 | **Roseburia sp. CAG:182** | 42 | 317 |
| 313 | **Parabacteroides sp. merdae-related_45_40** | 41 | 10 |
| 314 | **butyrate-producing bacterium SS3/4** | 41 | 248 |
| 315 | **Collinsella vaginalis** | 41 | 8 |
| 316 | **Faecalibacterium sp. CAG:82-related_59_9** | 41 | 99 |
| 317 | **Acinetobacter schindleri** | 40 | 0 |
| 318 | **Bacteroides oleiciplenus** | 40 | 55 |
| 319 | **Megamonas rupellensis** | 40 | 35 |
| 320 | **Blautia sp. Marseille-P2398** | 40 | 44 |
| 321 | **Clostridium sp. CAG:81** | 40 | 238 |
| 322 | **Comamonas kerstersii** | 40 | 0 |
| 323 | **Eggerthella sp. 1_3_56FAA** | 39 | 0 |
| 324 | **Enterobacter hormaechei** | 39 | 0 |
| 325 | **Rummeliibacillus stabekisii** | 39 | 0 |
| 326 | **Acinetobacter bereziniae** | 39 | 0 |
| 327 | **Lactobacillus pentosus** | 39 | 0 |
| 328 | **Eubacterium eligens CAG:72** | 39 | 102 |
| 329 | **Oscillibacter valericigenes** | 38 | 58 |
| 330 | **Bacteroides nordii** | 38 | 0 |
| 331 | **Clostridium sp. CAG:127** | 37 | 628 |
| 332 | **Eggerthella sp. HGA1** | 37 | 0 |
| 333 | **[Ruminococcus] gnavus** | 37 | 96 |
| 334 | **Ruminococcus sp. CAG:108** | 37 | 100 |
| 335 | **Olsenella mediterranea** | 37 | 0 |
| 336 | **Oscillibacter sp. PC13** | 37 | 56 |
| 337 | **Subdoligranulum sp. CAG:314** | 37 | 36 |
| 338 | **Oscillibacter ruminantium** | 37 | 68 |
| 339 | **Roseburia inulinivorans CAG:15** | 37 | 161 |
| 340 | **Finegoldia magna** | 36 | 0 |
| 341 | **Bacteroides sp. 3_1_23** | 36 | 42 |
| 342 | **Peptococcus niger** | 35 | 0 |
| 343 | **Bacteroides sp. HPS0048** | 35 | 0 |
| 344 | **Oribacterium sp. WCC10** | 35 | 15 |
| 345 | **Firmicutes bacterium CAG:552_39_19** | 34 | 68 |
| 346 | **Paraprevotella clara CAG:116** | 34 | 0 |
| 347 | **Clostridiales bacterium 41_21_two_genomes** | 34 | 338 |
| 348 | **Prevotella sp. CAG:604** | 34 | 247 |
| 349 | **Intestinimonas massiliensis** | 34 | 34 |
| 350 | **Clostridium sp. L2-50** | 34 | 123 |
| 351 | **Clostridium sp. CAG:1000** | 34 | 0 |
| 352 | **Firmicutes bacterium CAG:24** | 33 | 9 |
| 353 | **Enterococcus sp. FDAARGOS_375** | 33 | 0 |
| 354 | **Prevotella sp. CAG:891** | 33 | 34 |
| 355 | **Parabacteroides sp. D13** | 33 | 60 |
| 356 | **Enterococcus sp. HMSC05C03** | 33 | 0 |
| 357 | **Eubacterium siraeum CAG:80** | 33 | 39 |
| 358 | **Megasphaera sp. BL7** | 33 | 74 |
| 359 | **Bacteroides sp. D22** | 33 | 41 |
| 360 | **Lysinibacillus macroides** | 32 | 0 |
| 361 | **Klebsiella sp. MS 92-3** | 32 | 0 |
| 362 | **Sporobacter termitidis** | 32 | 36 |
| 363 | **Acinetobacter sp. NIPH 899** | 32 | 0 |
| 364 | **Pseudoflavonifractor capillosus** | 31 | 31 |
| 365 | **Lysinibacillus fusiformis** | 31 | 0 |
| 366 | **Peptoniphilus grossensis** | 31 | 0 |
| 367 | **Firmicutes bacterium CAG:102** | 31 | 206 |
| 368 | **Ruminococcus albus** | 31 | 33 |
| 369 | **Cutaneotrichosporon oleaginosum** | 31 | 0 |
| 370 | **Firmicutes bacterium HGW-Firmicutes-21** | 31 | 0 |
| 371 | **Marvinbryantia formatexigens** | 31 | 28 |
| 372 | **Collinsella bouchesdurhonensis** | 31 | 0 |
| 373 | **Acidovorax sp. 12322-1** | 31 | 0 |
| 374 | **Firmicutes bacterium CAG:321_26_22** | 31 | 51 |
| 375 | **Corallococcus sp. CAG:1435** | 30 | 143 |
| 376 | **Bacteroides sp. 3_1_40A** | 30 | 17 |
| 377 | **Ruminococcaceae bacterium D5** | 30 | 31 |
| 378 | **Bacteroidales bacterium 43_8** | 30 | 0 |
| 379 | **Anaerotruncus rubiinfantis** | 30 | 14 |
| 380 | **Ruminococcus champanellensis** | 30 | 57 |
| 381 | **Clostridium sp. CAG:349_48_7** | 30 | 25 |
| 382 | **Eubacterium rectale CAG:36** | 29 | 95 |
| 383 | **Clostridium sp. CAG:91** | 29 | 107 |
| 384 | **Butyricimonas sp. An62** | 29 | 0 |
| 385 | **Clostridiales bacterium GWF2_36_10** | 29 | 0 |
| 386 | **Alistipes finegoldii CAG:68** | 29 | 0 |
| 387 | **Flavonifractor sp. An10** | 28 | 30 |
| 388 | **Lactobacillus salivarius** | 28 | 0 |
| 389 | **Butyricimonas synergistica** | 28 | 0 |
| 390 | **Firmicutes bacterium CAG:65** | 28 | 160 |
| 391 | **Prevotella sp. CAG:5226** | 28 | 188 |
| 392 | **Romboutsia ilealis** | 27 | 8 |
| 393 | **Eubacterium sp. CAG:146** | 27 | 27 |
| 394 | **Clostridium sp. CAG:609** | 27 | 19 |
| 395 | **Blautia schinkii** | 27 | 0 |
| 396 | **Methanosphaera stadtmanae** | 27 | 0 |
| 397 | **Alistipes timonensis** | 26 | 0 |
| 398 | **Lysinibacillus sp. ZYM-1** | 26 | 0 |
| 399 | **Lachnospiraceae bacterium JC7** | 26 | 17 |
| 400 | **Bacteroides sp. CAG:709** | 26 | 140 |
| 401 | **Terrisporobacter glycolicus** | 26 | 0 |
| 402 | **Bacteroides sp. 3_1_19** | 25 | 22 |
| 403 | **bacterium LF-3** | 25 | 20 |
| 404 | **Bacteroides stercorirosoris** | 24 | 31 |
| 405 | **Peptoniphilus phoceensis** | 24 | 0 |
| 406 | **Oribacterium sp. P6A1** | 24 | 0 |
| 407 | **Prevotellamassilia timonensis** | 24 | 158 |
| 408 | **Ruminococcus faecis** | 24 | 10 |
| 409 | **Anaerofilum sp. An201** | 24 | 35 |
| 410 | **Dorea sp. 42_8** | 24 | 17 |
| 411 | **Eubacterium sp. CAG:251** | 24 | 290 |
| 412 | **Clostridium sp. CAG:389** | 23 | 45 |
| 413 | **Blautia sp. SF-50** | 23 | 9 |
| 414 | **Eubacterium sp. CAG:76** | 23 | 76 |
| 415 | **Clostridiales bacterium NK3B98** | 23 | 33 |
| 416 | **Firmicutes bacterium ASF500** | 23 | 46 |
| 417 | **Bacteroides bouchesdurhonensis** | 23 | 0 |
| 418 | **Eubacterium sp. 38_16** | 22 | 22 |
| 419 | **Clostridium sp. HGF2** | 22 | 0 |
| 420 | **Bacteroides sp. 9_1_42FAA** | 22 | 16 |
| 421 | **Mitsuokella jalaludinii** | 22 | 280 |
| 422 | **Paraprevotella clara** | 22 | 0 |
| 423 | **Blautia massiliensis** | 22 | 7 |
| 424 | **Coriobacteriaceae bacterium 68-1-3** | 22 | 0 |
| 425 | **Bacteroides sp. 43_108** | 22 | 22 |
| 426 | **Olsenella sp. An270** | 22 | 0 |
| 427 | **Isoptericola variabilis** | 22 | 0 |
| 428 | **Clostridium sp. KNHs209** | 21 | 13 |
| 429 | **Streptococcus parasanguinis** | 21 | 61 |
| 430 | **Gordonibacter massiliensis** | 21 | 0 |
| 431 | **Bilophila wadsworthia** | 21 | 436 |
| 432 | **uncultured Bacteroides sp.** | 21 | 46 |
| 433 | **Firmicutes bacterium CAG:552** | 21 | 31 |
| 434 | **Clostridium sp. SS2/1** | 21 | 60 |
| 435 | **Parabacteroides johnsonii** | 21 | 8 |
| 436 | **Flavonifractor sp. An100** | 20 | 50 |
| 437 | **Propionibacterium acidifaciens** | 20 | 0 |
| 438 | **Clostridium sp. CAG:914** | 20 | 29 |
| 439 | **Clostridium sp. 26_22** | 20 | 0 |
| 440 | **Bacteroides cellulosilyticus** | 20 | 82 |
| 441 | **Ruminococcaceae bacterium D16** | 20 | 163 |
| 442 | **Butyricicoccus pullicaecorum** | 20 | 30 |
| 443 | **Actinomyces sp. ICM47** | 20 | 0 |
| 444 | **Olsenella sp. Marseille-P2300** | 20 | 0 |
| 445 | **Collinsella tanakaei** | 20 | 15 |
| 446 | **Gemmiger sp. An120** | 19 | 39 |
| 447 | **Pseudoflavonifractor sp. An184** | 19 | 35 |
| 448 | **Peptoniphilus sp. HMSC062D09** | 19 | 0 |
| 449 | **Bacteroides sp. 3_1_13** | 19 | 36 |
| 450 | **Flavonifractor sp. An306** | 19 | 32 |
| 451 | **Firmicutes bacterium CAG:145** | 19 | 15 |
| 452 | **Peptoniphilus timonensis** | 18 | 0 |
| 453 | **Eubacterium ventriosum** | 18 | 70 |
| 454 | **Clostridiales bacterium 43-6** | 18 | 0 |
| 455 | **Cloacibacillus evryensis** | 18 | 0 |
| 456 | **Akkermansia muciniphila CAG:154** | 18 | 13 |
| 457 | **Oribacterium sp. C9** | 18 | 0 |
| 458 | **Olsenella uli** | 18 | 11 |
| 459 | **Prevotella sp. CAG:1092** | 17 | 299 |
| 460 | **Slackia heliotrinireducens** | 17 | 0 |
| 461 | **Lagierella massiliensis** | 17 | 0 |
| 462 | **Firmicutes bacterium CAG:194** | 17 | 14 |
| 463 | **Lachnospiraceae bacterium 28-4** | 17 | 0 |
| 464 | **Oribacterium sp. NK2B42** | 17 | 0 |
| 465 | **Bacteroides eggerthii CAG:109** | 17 | 0 |
| 466 | **Eggerthella timonensis** | 17 | 0 |
| 467 | **Clostridium sp. CAG:762** | 17 | 79 |
| 468 | **Brachyspira sp. CAG:484** | 17 | 91 |
| 469 | **methanogenic archaeon mixed culture ISO4-G1** | 17 | 38 |
| 470 | **Eggerthella sp. 51_9** | 17 | 0 |
| 471 | **Ruminococcus sp. CAG:379** | 17 | 27 |
| 472 | **Bacteroides timonensis** | 17 | 98 |
| 473 | **Oscillibacter sp. CAG:155** | 17 | 36 |
| 474 | **Peptoniphilus senegalensis** | 17 | 0 |
| 475 | **Lachnospira pectinoschiza** | 16 | 117 |
| 476 | **Firmicutes bacterium CAG:95** | 16 | 197 |
| 477 | **Megamonas sp. Calf98-2** | 16 | 10 |
| 478 | **Massilimaliae massiliensis** | 16 | 14 |
| 479 | **Anaeromassilibacillus sp. An200** | 16 | 17 |
| 480 | **Odoribacter sp. 43_10** | 16 | 0 |
| 481 | **Bacteroides mediterraneensis** | 16 | 11 |
| 482 | **Bacteroides dorei CAG:222** | 16 | 7 |
| 483 | **Bacteroides sp. 4_3_47FAA** | 16 | 13 |
| 484 | **Firmicutes bacterium CAG:534** | 16 | 314 |
| 485 | **Ruminococcaceae bacterium FB2012** | 16 | 0 |
| 486 | **Clostridium disporicum** | 16 | 0 |
| 487 | **Neglecta timonensis** | 16 | 13 |
| 488 | **Bacteroides uniformis CAG:3** | 15 | 9 |
| 489 | **Clostridium bolteae CAG:59** | 15 | 0 |
| 490 | **Agathobaculum desmolans** | 15 | 61 |
| 491 | **Bacteroides sartorii** | 15 | 5 |
| 492 | **Bacteroides sp. D2** | 15 | 63 |
| 493 | **Lachnoclostridium sp. An196** | 15 | 151 |
| 494 | **Acidaminococcus massiliensis** | 15 | 0 |
| 495 | **Raoultibacter massiliensis** | 15 | 0 |
| 496 | **Clostridium sp. SN20** | 15 | 0 |
| 497 | **Eggerthella sp. YY7918** | 15 | 0 |
| 498 | **Bacteroides sp. 43_46** | 15 | 0 |
| 499 | **Lachnospiraceae bacterium 7_1_58FAA** | 15 | 25 |
| 500 | **Eggerthellaceae bacterium AT8** | 15 | 0 |
| 501 | **Alistipes sp. AL-1** | 15 | 10 |
| 502 | **Ruminococcus sp. SR1/5** | 15 | 0 |
| 503 | **Bacteroides fragilis CAG:558** | 15 | 0 |
| 504 | **[Clostridium] thermosuccinogenes** | 14 | 0 |
| 505 | **Hydrogenoanaerobacterium saccharovorans** | 14 | 14 |
| 506 | **Alistipes sp. 58_9_plus** | 14 | 0 |
| 507 | **Eubacterium sp. CAG:86** | 14 | 92 |
| 508 | **Faecalibacterium sp. CAG:1138** | 14 | 29 |
| 509 | **Actinomyces oris** | 14 | 0 |
| 510 | **Dorea sp. CAG:105** | 14 | 0 |
| 511 | **Eubacteriaceae bacterium CHKCI005** | 14 | 17 |
| 512 | **Succinivibrio dextrinosolvens** | 14 | 32 |
| 513 | **Lactococcus garvieae** | 14 | 0 |
| 514 | **Bacteroides sp. 1_1_30** | 14 | 33 |
| 515 | **[Clostridium] asparagiforme** | 14 | 9 |
| 516 | **Ruminococcus sp. CAG:9-related_41_34** | 14 | 13 |
| 517 | **Enterococcus sp. HMSC072H05** | 14 | 0 |
| 518 | **Firmicutes bacterium CAG:475** | 14 | 39 |
| 519 | **Clostridium sp. CAG:440** | 14 | 0 |
| 520 | **Coprobacillus sp. 8_1_38FAA** | 14 | 35 |
| 521 | **Alloprevotella rava** | 14 | 14 |
| 522 | **Collinsella ihuae** | 13 | 0 |
| 523 | **Coprobacillus sp. CAG:235_29_27** | 13 | 0 |
| 524 | **Megasphaera sp. NM10** | 13 | 39 |
| 525 | **Enterorhabdus caecimuris** | 13 | 21 |
| 526 | **Peptoniphilus harei** | 13 | 0 |
| 527 | **Prevotella sp. CAG:755** | 13 | 13 |
| 528 | **Ruminococcus sp. CAG:579** | 13 | 0 |
| 529 | **Paraprevotella xylaniphila** | 13 | 0 |
| 530 | **Enterococcus sp. 5B7_DIV0075** | 12 | 0 |
| 531 | **Tyzzerella nexilis** | 12 | 81 |
| 532 | **Ruminococcus sp. CAG:57** | 12 | 36 |
| 533 | **Parabacteroides sp. Marseille-P3763** | 12 | 0 |
| 534 | **Raoultibacter timonensis** | 12 | 0 |
| 535 | **Bacteroides thetaiotaomicron CAG:40** | 12 | 0 |
| 536 | **Clostridiales bacterium SK-Y3** | 12 | 0 |
| 537 | **Eubacterium sp. 41_20** | 12 | 62 |
| 538 | **Mobilibacterium timonense** | 12 | 0 |
| 539 | **Arabia massiliensis** | 12 | 0 |
| 540 | **Bacteroides vulgatus CAG:6** | 12 | 7 |
| 541 | **Lysinibacillus boronitolerans** | 12 | 0 |
| 542 | **Ruminococcus sp. CAG:108-related_41_35** | 11 | 50 |
| 543 | **Acinetobacter sp. LCT-H3** | 11 | 0 |
| 544 | **Odoribacter splanchnicus CAG:14** | 11 | 0 |
| 545 | **Synergistes jonesii** | 11 | 0 |
| 546 | **Olsenella profusa** | 11 | 6 |
| 547 | **Synergistes sp. 3_1_syn1** | 11 | 0 |
| 548 | **Bilophila sp. 4_1_30** | 11 | 78 |
| 549 | **Clostridium sp. CAG:798** | 11 | 61 |
| 550 | **Bacteroides sp. 2_1_33B** | 11 | 0 |
| 551 | **Prevotella sp. 885** | 11 | 146 |
| 552 | **Intestinibacter bartlettii** | 11 | 13 |
| 553 | **Enterorhabdus mucosicola** | 10 | 11 |
| 554 | **Bacteroides cellulosilyticus CAG:158** | 10 | 30 |
| 555 | **Enterococcus sp. 3H8_DIV0648** | 10 | 0 |
| 556 | **Clostridium sp. CAG:269** | 10 | 115 |
| 557 | **Candida parapsilosis** | 10 | 0 |
| 558 | **Clostridium nexile CAG:348** | 10 | 74 |
| 559 | **Weissella sp. DD23** | 10 | 0 |
| 560 | **Actinomyces dentalis** | 9 | 0 |
| 561 | **uncultured crAssphage** | 9 | 9 |
| 562 | **Tissierellia bacterium S5-A11** | 9 | 0 |
| 563 | **Clostridium sp. ASBs410** | 9 | 0 |
| 564 | **Roseburia sp. CAG:471** | 9 | 119 |
| 565 | **Paeniclostridium sordellii** | 9 | 0 |
| 566 | **Collinsella stercoris** | 9 | 0 |
| 567 | **Bacteroides coprophilus** | 9 | 8 |
| 568 | **Bacteroides sp. 3_1_33FAA** | 9 | 17 |
| 569 | **Enterococcus gilvus** | 9 | 0 |
| 570 | **Acidaminococcus sp. CAG:542** | 8 | 0 |
| 571 | **Alistipes sp. CAG:29** | 8 | 30 |
| 572 | **Prevotella sp. CAG:617** | 8 | 8 |
| 573 | **Leuconostoc lactis** | 8 | 0 |
| 574 | **Collinsella sp. An2** | 8 | 0 |
| 575 | **Actinomyces sp. oral taxon 175** | 8 | 0 |
| 576 | **Clostridiales bacterium VE202-21** | 8 | 0 |
| 577 | **Listeria monocytogenes** | 8 | 0 |
| 578 | **Clostridium sp. 7_2_43FAA** | 8 | 0 |
| 579 | **Paraclostridium bifermentans** | 8 | 0 |
| 580 | **Enterococcus pallens** | 8 | 0 |
| 581 | **[Desulfotomaculum] guttoideum** | 7 | 0 |
| 582 | **Anaerococcus prevotii** | 7 | 0 |
| 583 | **Actinomyces viscosus** | 7 | 0 |
| 584 | **Bacteroides finegoldii** | 7 | 33 |
| 585 | **Clostridium celatum** | 7 | 0 |
| 586 | **Bacteroides stercoris CAG:120** | 7 | 0 |
| 587 | **Enterococcus saccharolyticus** | 7 | 0 |
| 588 | **Enterococcus malodoratus** | 7 | 0 |
| 589 | **Clostridium sp. CAG:470** | 7 | 10 |
| 590 | **Actinomyces odontolyticus** | 7 | 0 |
| 591 | **Peptoniphilus duerdenii** | 7 | 0 |
| 592 | **Actinomyces sp. ICM58** | 7 | 0 |
| 593 | **Ruminococcus sp. 37_24** | 7 | 0 |
| 594 | **Alistipes putredinis CAG:67** | 7 | 9 |
| 595 | **Fusobacterium mortiferum** | 7 | 0 |
| 596 | **Collinsella phocaeensis** | 7 | 0 |
| 597 | **Dialister succinatiphilus** | 7 | 196 |
| 598 | **Enterococcus sp. kppr-6** | 6 | 0 |
| 599 | **Eubacterium callanderi** | 6 | 0 |
| 600 | **Sutterella wadsworthensis CAG:135** | 6 | 18 |
| 601 | **Clostridiales bacterium 36_14** | 6 | 52 |
| 602 | **Prevotella lascolaii** | 6 | 11 |
| 603 | **Collinsella intestinalis** | 6 | 0 |
| 604 | **Lachnospiraceae bacterium 5_1_63FAA** | 6 | 33 |
| 605 | **Actinomyces sp. ICM39** | 6 | 0 |
| 606 | **Acinetobacter sp. CIP 101934** | 6 | 0 |
| 607 | **Acinetobacter lwoffii** | 5 | 0 |
| 608 | **Peptoniphilus sp. BV3AC2** | 5 | 0 |
| 609 | **Bifidobacterium bifidum CAG:234** | 5 | 0 |
| 610 | **Bifidobacterium pseudocatenulatum CAG:263** | 5 | 0 |
| 611 | **Lactobacillus rhamnosus** | 5 | 0 |
| 612 | **Prevotella sp. CAG:732** | 5 | 65 |
| 613 | **Megamonas funiformis CAG:377** | 5 | 0 |
| 614 | **Megamonas hypermegale** | 4 | 124 |
| 615 | **Eubacterium sp. CAG76_36_125** | 4 | 30 |
| 616 | **Bifidobacterium adolescentis CAG:119** | 4 | 18 |
| 617 | **Roseburia sp. CAG:197** | 0 | 47 |
| 618 | **Bacteroides sp. CAG:98** | 0 | 41 |
| 619 | **Parasutterella excrementihominis CAG:233** | 0 | 48 |
| 620 | **Clostridium sp. CAG:122** | 0 | 98 |
| 621 | **Firmicutes bacterium CAG:313** | 0 | 124 |
| 622 | **Clostridiales bacterium KLE1615** | 0 | 301 |
| 623 | **Elusimicrobium sp. An273** | 0 | 124 |
| 624 | **Bifidobacterium dentium** | 0 | 5 |
| 625 | **Clostridia bacterium UC5.1-1D1** | 0 | 8 |
| 626 | **Tyzzerella sp. Marseille-P3062** | 0 | 35 |
| 627 | **Acidaminococcus intestini** | 0 | 7 |
| 628 | **Prevotella sp. CAG:386** | 0 | 64 |
| 629 | **uncultured Lachnospira sp.** | 0 | 248 |
| 630 | **Clostridium sp. M62/1** | 0 | 10 |
| 631 | **Sellimonas intestinalis** | 0 | 23 |
| 632 | **Clostridium sp. CAG:780** | 0 | 114 |
| 633 | **Clostridium sp. AT4** | 0 | 26 |
| 634 | **Roseburia sp. CAG:380** | 0 | 16 |
| 635 | **Ruminococcus sp. CAG:330** | 0 | 23 |
| 636 | **Clostridium sp. CAG:575** | 0 | 4 |
| 637 | **Clostridium sp. CAG:62** | 0 | 74 |
| 638 | **Lachnospiraceae bacterium 8_1_57FAA** | 0 | 11 |
| 639 | **Eubacterium sp. CAG:603** | 0 | 22 |
| 640 | **Ruminococcaceae bacterium cv2** | 0 | 15 |
| 641 | **Akkermansia sp. CAG:344** | 0 | 58 |
| 642 | **Clostridium sp. 44_14** | 0 | 54 |
| 643 | **Clostridium sp. CAG:628** | 0 | 107 |
| 644 | **Ruminococcus sp. CAG:624** | 0 | 32 |
| 645 | **Roseburia sp. CAG:303** | 0 | 451 |
| 646 | **Lactobacillus rogosae** | 0 | 33 |
| 647 | **Roseburia sp. CAG:309** | 0 | 25 |
| 648 | **Acholeplasma sp. CAG:878** | 0 | 115 |
| 649 | **Shigella sonnei** | 0 | 35 |
| 650 | **Megasphaera elsdenii CAG:570** | 0 | 7 |
| 651 | **Fusobacterium sp. CAG:439** | 0 | 8 |
| 652 | **Bifidobacterium pseudolongum** | 0 | 9 |
| 653 | **Bacteroides sp. CAG:770** | 0 | 38 |
| 654 | **Bacteroides sp. 14(A)** | 0 | 11 |
| 655 | **Anaerovorax odorimutans** | 0 | 15 |
| 656 | **Bifidobacterium merycicum** | 0 | 7 |
| 657 | **Ruminococcus callidus** | 0 | 268 |
| 658 | **Eubacterium sp. CAG:252** | 0 | 27 |
| 659 | **Clostridium sp. CAG:253** | 0 | 42 |
| 660 | **Clostridium sp. ATCC BAA-442** | 0 | 20 |
| 661 | **Coprobacillus sp. 28_7** | 0 | 21 |
| 662 | **Clostridium sp. SCN 57-10** | 0 | 40 |
| 663 | **Ruminococcus sp. CAG:403** | 0 | 7 |
| 664 | **Clostridium sp. 42_12** | 0 | 14 |
| 665 | **Firmicutes bacterium CAG:65_45_313** | 0 | 108 |
| 666 | **Lachnospiraceae bacterium CAG:25** | 0 | 8 |
| 667 | **Sutterella parvirubra** | 0 | 15 |
| 668 | **Eubacterium sp. 45_250** | 0 | 17 |
| 669 | **Firmicutes bacterium CAG:272_52_7** | 0 | 26 |
| 670 | **Clostridium sp. 29_15** | 0 | 18 |
| 671 | **Veillonella dispar** | 0 | 94 |
| 672 | **Bacteroides sp. CAG:530** | 0 | 145 |
| 673 | **Flavonifractor sp. An82** | 0 | 15 |
| 674 | **Firmicutes bacterium CAG:449** | 0 | 81 |
| 675 | **Eubacterium ramulus** | 0 | 27 |
| 676 | **Clostridium sp. CAG:75** | 0 | 26 |
| 677 | **Shigella flexneri** | 0 | 10 |
| 678 | **Coprococcus comes CAG:19** | 0 | 9 |
| 679 | **Firmicutes bacterium CAG:341** | 0 | 52 |
| 680 | **Candidatus Gastranaerophilales bacterium HUM_1** | 0 | 102 |
| 681 | **Clostridium sp. CAG:813** | 0 | 49 |
| 682 | **Proteobacteria bacterium CAG:495** | 0 | 31 |
| 683 | **Coprobacillus sp. CAG:698** | 0 | 130 |
| 684 | **Prevotella stercorea** | 0 | 117 |
| 685 | **Gemmiger sp. An50** | 0 | 16 |
| 686 | **Clostridium sp. 26_21** | 0 | 12 |
| 687 | **Clostridium sp. CAG:217** | 0 | 20 |
| 688 | **Butyrivibrio crossotus** | 0 | 11 |
| 689 | **Coprococcus sp. CAG:782** | 0 | 11 |
| 690 | **Prevotella sp. P3-122** | 0 | 18 |
| 691 | **Blautia sp. CAG:52** | 0 | 31 |
| 692 | **uncultured organism** | 0 | 7 |
| 693 | **Roseburia sp. 499** | 0 | 46 |
| 694 | **Clostridium sp. CAG:277** | 0 | 170 |
| 695 | **Mitsuokella multacida** | 0 | 126 |
| 696 | **Azospirillum sp. 51_20** | 0 | 8 |
| 697 | **Ruminococcus sp. DSM 100440** | 0 | 37 |
| 698 | **Clostridium sp. CAG:62_40_43** | 0 | 30 |
| 699 | **Butyrivibrio crossotus CAG:259** | 0 | 10 |
| 700 | **Azospirillum sp. CAG:239** | 0 | 82 |
| 701 | **Prevotella sp. CAG:520** | 0 | 540 |
| 702 | **Lachnospiraceae bacterium TF01-11** | 0 | 111 |
| 703 | **Burkholderiales bacterium 1_1_47** | 0 | 118 |
| 704 | **Bacteroides massiliensis** | 0 | 134 |
| 705 | **Eubacterium sp. CAG:248** | 0 | 294 |
| 706 | **Clostridiales bacterium VE202-14** | 0 | 17 |
| 707 | **Eubacterium sp. 36_13** | 0 | 36 |
| 708 | **Veillonella atypica** | 0 | 41 |
| 709 | **Sutterella sp. 54_7** | 0 | 5 |
| 710 | **Dakarella massiliensis** | 0 | 64 |
| 711 | **Clostridium ventriculi** | 0 | 119 |
| 712 | **Sutterella sp. CAG:351** | 0 | 44 |
| 713 | **Bacteroides acidifaciens** | 0 | 9 |
| 714 | **Lachnospiraceae bacterium KHCPX20** | 0 | 7 |
| 715 | **Roseburia sp. CAG:10041_57** | 0 | 38 |
| 716 | **Veillonella sp. DORA_A_3_16_22** | 0 | 33 |
| 717 | **Selenomonas ruminantium** | 0 | 6 |
| 718 | **Emergencia timonensis** | 0 | 14 |
| 719 | **Roseburia sp. CAG:45** | 0 | 62 |
| 720 | **Azospirillum sp. 47_25** | 0 | 8 |
| 721 | **Fournierella massiliensis** | 0 | 23 |
| 722 | **Tenericutes bacterium HGW-Tenericutes-4** | 0 | 16 |
| 723 | **Blastocystis sp. subtype 1** | 0 | 33 |
| 724 | **Prevotella bryantii** | 0 | 8 |
| 725 | **Roseburia sp. 831b** | 0 | 46 |
| 726 | **Bacteroides sp. CAG:754** | 0 | 25 |
| 727 | **Roseburia sp. 40_7** | 0 | 11 |
| 728 | **Roseburia sp. CAG:100** | 0 | 70 |
| 729 | **Bacteroides congonensis** | 0 | 14 |
| 730 | **Clostridium sp. CAG:12237_41** | 0 | 36 |
| 731 | **Lachnospiraceae bacterium 10-1** | 0 | 20 |
| 732 | **Eubacterium sp. CAG:38** | 0 | 452 |
| 733 | **Azospirillum sp. CAG:260** | 0 | 16 |
| 734 | **Bacteroides ovatus CAG:22** | 0 | 36 |
| 735 | **Firmicutes bacterium CAG_194_44_15** | 0 | 7 |
| 736 | **Veillonella parvula** | 0 | 14 |
| 737 | **Clostridium sp. CAG:265** | 0 | 41 |
| 738 | **Coprococcus sp. ART55/1** | 0 | 9 |
| 739 | **Parasutterella excrementihominis** | 0 | 63 |
| 740 | **Roseburia intestinalis CAG:13** | 0 | 190 |
| 741 | **Roseburia sp. CAG:50** | 0 | 13 |
| 742 | **uncultured Roseburia sp.** | 0 | 36 |
| 743 | **Akkermansia glycaniphila** | 0 | 18 |
| 744 | **Mycoplasma sp. CAG:611** | 0 | 61 |
| 745 | **Eggerthella sp. CAG:298** | 0 | 43 |
| 746 | **Clostridium sp. CAG:632** | 0 | 226 |
| 747 | **Prevotella ruminicola** | 0 | 7 |
| 748 | **Drancourtella massiliensis** | 0 | 18 |
| 749 | **Lachnoclostridium sp. An14** | 0 | 19 |
| 750 | **Eubacterium sp. CAG:115** | 0 | 14 |
| 751 | **Bacteroides sp. CAG:1076** | 0 | 4 |
| 752 | **Coprococcus sp. CAG:131** | 0 | 26 |
| 753 | **Veillonella sp. oral taxon 158** | 0 | 12 |
| 754 | **Phascolarctobacterium sp. CAG:266** | 0 | 14 |
| 755 | **Lachnoclostridium edouardi** | 0 | 11 |
| 756 | **Prevotella stercorea CAG:629** | 0 | 24 |
| 757 | **Alloprevotella tannerae** | 0 | 7 |
| 758 | **Olsenella sp. oral taxon 807** | 0 | 8 |
